# Uncertainty-aware longitudinal prediction of multiple adverse pregnancy outcomes

**DOI:** 10.64898/2026.11.15.26368140

**Authors:** Yanqi Wu, Sima Asvadi, Myrthe van der Ven, Elisabetta Peri, Mengzhu Xu, M. Beatrijs van der Hout-van der Jagt, Judith OEH van Laar, Chen Chen, Qiong Luo, Massimo Mischi, S. Guid Oei, Xi Long

## Abstract

**Background:** Risk of pregnancy complications is reassessed at each antenatal visit, but whether earlier predictions add information beyond the most recent one is unclear. Classifying every prediction as low or high risk can leave uncertainty unexpressed. Whether reliability per visit holds across visits is also unclear.

**Methods:** This secondary analysis included 9,289 nulliparous women from the nuMoM2b cohort (eight US centres, 2010–13). Logistic regression models predicted gestational diabetes, pre-eclampsia, preterm birth, suspected fetal growth restriction and large for gestational age at 6–13, 16–21 and 22–29 weeks (Visits 1–3). Mondrian conformal prediction reported ‘no’ (low risk), ‘yes’ (high risk), or both (review). To test whether prediction history adds information beyond the Visit 3 prediction, we compared, among women classified as high risk at Visit 3, those first classified as high risk at Visit 2 or 3 (emerging high) with those classified as high risk at all three visits (persistent high), adjusting for Visit 3 predicted probability.

**Findings:** The adjusted proportion with the outcome was higher in the emerging-high than the persistent-high group, by 6.1 percentage points for large for gestational age (95% CI −0.4 to 12.2) and 3.8 percentage points for suspected fetal growth restriction (−0.9 to 8.1). Other outcomes showed adjusted differences close to zero. With conformal prediction calibrated to a 90% level at each visit, the reported answers included the actual outcome in 89.6–90.6% of women at each visit (coverage). Coverage decreased when each woman’s three visits were considered together (79.0–85.4%).

**Interpretation:** For fetal growth outcomes, how a woman’s predicted risk evolved during pregnancy may add information beyond her Visit 3 prediction. For repeated clinical use, the target coverage level should reflect the acceptable risk of incorrect low- or high-risk classifications over a woman’s follow-up and the clinical decisions supported at each visit.

**Funding:** None.

**Research in context:** *Evidence before this study:* We searched PubMed from database inception to Oct 28, 2026, for English-language studies in humans, combining terms for gestational diabetes, pre-eclampsia, preterm birth, fetal growth restriction or small for gestational age, and large for gestational age with terms for prediction models and for longitudinal or repeated assessment, risk trajectories, conformal prediction, or predictive uncertainty. We also searched for conformal prediction applied to longitudinal clinical data. Most models predicted a single complication at a fixed gestational age. Longitudinal models updated risk using repeated measurements of blood pressure, biomarkers or fetal biometry, and some showed that changes in these measurements, including fetal growth velocity, improved prediction. We found no study that tested whether a woman’s earlier predictions add information beyond her most recent prediction. Individual-level uncertainty was rarely quantified in antenatal prediction, and we found no application of conformal prediction in antenatal care. Conformal methods have been used for repeated prediction in other clinical settings, but we found no evaluation of how often prediction sets include the actual outcome across a patient’s successive predictions.

## Introduction

Adverse pregnancy outcomes are important causes of maternal and neonatal morbidity and mortality^1–3^. These outcomes include gestational diabetes mellitus (GDM), pre-eclampsia (PE), preterm birth (PTB), fetal growth restriction (FGR) and large for gestational age (LGA). Assessing the risk of these outcomes is part of routine antenatal care. Antenatal guidelines recommend repeating the assessment at every routine appointment and reviewing the plan of care each time, because a woman’s risk of developing a pregnancy complication can change as pregnancy progresses^4,5^. Later visits provide additional maternal measurements and information about fetal development, allowing risk estimates to be updated^6^.

Prediction models estimate the risk of specific adverse pregnancy outcomes at defined gestational ages^7,8^. A model developed for a later visit can combine information collected at earlier visits with newly available measurements, and dynamic prediction methods can update risk using repeated measurements^9–12^. Repeated assessment therefore produces a sequence of predictions for each woman. Two women with similar predicted risks later in pregnancy may have different prediction histories: one may have had an elevated risk estimate from early pregnancy, whereas the other may have had lower risk estimates at earlier visits. Because a later model can already incorporate earlier measurements, it remains unclear whether a woman’s earlier predictions add information beyond her current predicted risk.

To interpret these repeated predictions, it is also necessary to consider how each prediction is reported. Prediction models provide estimated probabilities^7,8^, which are often converted into low- or high-risk classifications using a single cut-off. This forces a classification on every woman, including those whose predicted probabilities fall in a range where women who will and will not develop the complication are difficult to tell apart^13^. Conformal prediction uses these predicted probabilities to report a set of possible answers rather than always a single category^14^. For example, for PE, this prediction set may contain ‘no PE’, ‘PE’, or both (‘no PE’ and ‘PE’). A set containing only ‘no PE’ is a low-risk prediction, and a set containing only ‘PE’ is a high-risk prediction. A set containing both is labelled review, meaning that conformal prediction did not give a low- or high-risk classification at that visit.

The conformal prediction method uses a separate group of women with known outcomes to decide which answers to include in each prediction set. This step is called calibration. Calibration aims to ensure that, for a specified proportion of women, such as 90%, the prediction set includes the actual outcome, provided that the women being assessed are similar to those used for calibration^14^. This proportion is called coverage. When the complication occurs in only a small proportion of women, coverage across all women may be close to the target even if coverage among women who develop the complication is much lower. To address this, Mondrian conformal prediction applies the coverage target separately to women who develop the complication and to those who do not^15^. Applied at each antenatal visit, Mondrian conformal prediction gives a low-risk, high-risk or review prediction, so that clinicians can follow how a woman’s predicted risk changes across pregnancy. The coverage target applies to each visit separately, so it does not establish how often the prediction sets include the actual outcome at every visit for the same woman.

A woman may also receive predictions for several different outcomes at the same visit. Because separate outcome-specific models can use the same clinical measurements, a change in those measurements may shift several predictions in the same or opposite directions. Examining the contributions of these shared predictors may help clinicians understand why predictions for different outcomes change together.

In this study, we developed models to predict five adverse pregnancy outcomes at three antenatal visits (6–13, 16–21 and 22–29 weeks of gestation) and used Mondrian conformal prediction to express each prediction as low risk, high risk or review. Among women classified the same way at Visit 3, we compared the proportion with the complication between groups with different prediction histories, after accounting for the Visit 3 predicted probability. We also examined whether predictions for different outcomes changed together, and which shared predictors contributed to these changes. Finally, we assessed how often the prediction sets included the actual outcome at each visit, and how often they did so at all three visits for the same woman.

## Methods

### Study population and data source

The Nulliparous Pregnancy Outcomes Study: Monitoring Mothers-to-Be (nuMoM2b) recruited nulliparous women with singleton pregnancies at eight US clinical centres between September 2010 and September 2013^16^. This secondary analysis used data from all 9,289 participants who consented to data sharing. The study included three scheduled antenatal research visits: Visit 1 at 6–13 weeks, Visit 2 at 16– 21 weeks and Visit 3 at 22–29 weeks of gestation, followed by a delivery assessment^16^. An overview of the study design is shown in Fig. 1. Participants were enrolled at Visit 1 and followed to delivery; no participants entered the cohort after enrolment. The present analysis was approved by the Eunice Kennedy Shriver National Institute of Child Health and Human Development review board (15 December 2021; request ID 11961) and the Eindhoven University of Technology ethical review board (1 November 2021; reference ERB2021BME4). The study was conducted in accordance with relevant ethical regulations. Baseline characteristics of the cohort are reported in Table 1.

**Figure 1.**
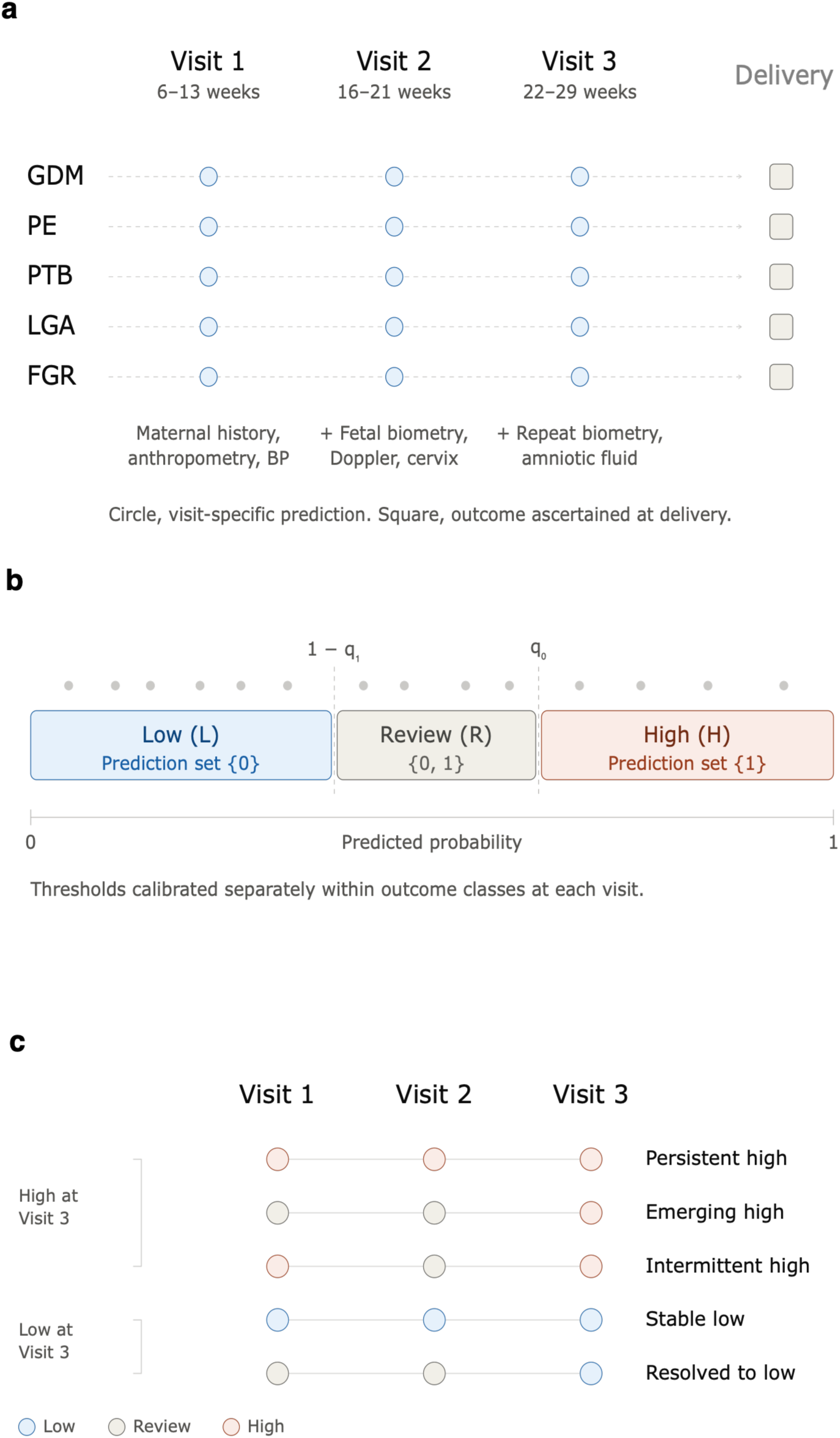
Study design and analytical overview of longitudinal uncertainty-aware prediction across gestation. a, outcome-specific predictions were generated at three antenatal visits as clinical and fetal information accumulated across gestation. Five adverse pregnancy outcomes were considered: gestational diabetes mellitus (GDM), pre-eclampsia (PE), preterm birth (PTB), large for gestational age (LGA), and fetal growth restriction (FGR). b, for each outcome and visit, binary conformal prediction was represented as an ordered partition of the predicted event probability into low-risk (L), review (R) and high-risk (H) states. The two thresholds were calibrated separately for each outcome and visit. c, visit-specific states were followed longitudinally to characterise prediction histories. Representative trajectories include persistent high, emerging high, intermittent high, stable low, and resolved-to-low patterns. Abbreviations: GDM, gestational diabetes mellitus; PE, pre-eclampsia; PTB, preterm birth; LGA, large for gestational age; FGR, fetal growth restriction; L, low risk; R, review; H, high risk.

**Table 1.** Baseline characteristics of the study cohort.

| **Characteristic** | **Overall cohort (N=9,289)** |
|---|---|
| Participants, n | 9,289 |
| Maternal age, years | 26.9 (5.7) |
| BMI at Visit 1, kg/m² | 26.4 (6.3) |
| **Race/ethnicity, n (%)** |  |
| Non-Hispanic White | 5,595 (60.3%) |
| Non-Hispanic Black | 1,248 (13.4%) |
| Hispanic | 1,599 (17.2%) |
| American Indian / Asian / Native Hawaiian / other | 405 (4.4%) |
| Other | 55 (0.6%) |
| Multiracial | 377 (4.1%) |
| **Education, n (%)** |  |
| Less than high school or high school | 1,850 (20.0%) |
| Some college or associate/technical degree | 2,764 (29.8%) |
| Completed college | 2,544 (27.4%) |
| Beyond college | 2,113 (22.8%) |
| Polycystic ovary syndrome, n (%) | 386 (4.4%) |
| Family history of diabetes, n (%) | 1,918 (22.2%) |
| Systolic blood pressure at Visit 1, mmHg | 109.2 (10.9) |
| Diastolic blood pressure at Visit 1, mmHg | 67.0 (8.4) |

**Table note**: Values are mean (SD) for continuous variables and n (%) for categorical variables. Percentages for categorical variables were calculated among participants with non-missing data. Baseline characteristics were summarised at Visit 1 for the full study cohort (N=9,289). Data were missing for 47 participants for maternal age, 210 for BMI, 10 for race/ethnicity, 18 for education, 508 for polycystic ovary syndrome, 663 for family history of diabetes, and 202 for each blood-pressure measure.

### Adverse pregnancy outcomes

Outcome data were abstracted from medical records by trained and certified research staff after delivery^16^. Gestational age, used in the definitions below, was established at enrolment (6–13 weeks) using standardised ultrasonography performed by certified sonographers^16^. GDM was ascertained by oral glucose tolerance testing at 24–28 weeks according to site protocols, using the nuMoM2b diagnostic criteria^17^ detailed in Supplementary Method S1. PE comprised mild, severe or superimposed pre-eclampsia and eclampsia, classified according to nuMoM2b study definitions after detailed chart review, with atypical cases classified by consensus of the principal investigators^16,17^. PTB was defined as delivery between 20 and 36 completed weeks of gestation, irrespective of indication^16^. FGR was identified using the clinical abstraction variable for suspected fetal growth restriction before delivery (nuMoM2b variable CMAE08)^16^. LGA was defined as birthweight at or above the 90th percentile according to sex-specific, first-born birthweight-for-gestational-age values from the 2017 US reference by Aris et al.^18^.

The number of eligible participants varied across outcomes, depending on the data needed to determine each outcome and on outcome-specific exclusions. Outcomes were modelled separately, and the same participant could be included in more than one outcome analysis. GDM required an evaluable pregnancy diabetes outcome, with pre-existing diabetes excluded; PTB required a valid gestational age at delivery; LGA required gestational age at delivery, birthweight, and fetal sex; and FGR required an available clinical abstraction label. PE status was available for all participants. Visit 3 analyses were restricted to pregnancies with delivery at or after 30+0 weeks, corresponding to the end of the Visit 3 assessment window. For FGR, Visit 3 predictions were excluded for participants whose suspected FGR was recorded on or before the date of their Visit 3 assessment (n = 28); their Visit 1 and Visit 2 predictions were retained. Routine GDM screening and diagnosis at 24–28 weeks overlap the Visit 3 window, so Visit 3 discrimination for GDM does not represent prediction ahead of diagnosis.

### Prediction models and conformal prediction

Candidate predictors were derived from variables used in previous nuMoM2b studies^10,19,20^ and were organised according to their availability at each antenatal visit. Three predictor sets were constructed for Visit 1 (6–13 weeks), Visit 2 (16–21 weeks), and Visit 3 (22–29 weeks), with information from earlier visits retained at subsequent visits. The predictor sets included maternal demographic and clinical characteristics, medical and obstetric history, anthropometric and blood-pressure measurements, lifestyle and dietary variables, psychological measures, routine laboratory measurements, and ultrasound-derived fetal measurements. Variables were excluded if they were not available by the corresponding visit or had more than 20% missing values. The same visit-specific candidate predictor set was used for all five outcomes. Detailed predictor definitions and availability are provided in Supplementary Table S1.

Separate prediction models were developed for each outcome at each antenatal visit using elastic-net logistic regression^21^, with hyperparameters selected within the training data of each iteration. Models were evaluated by participant-level stratified 10-fold cross-validation, with each fold held out in turn so that every participant received a prediction from a model fitted without their data. The same fold assignment was used at Visits 1 and 2, whereas Visit 3 was partitioned separately within the Visit 3 analysis set restricted to deliveries at or after 30+0 weeks. Discrimination, the ability of the predicted probabilities to separate participants with and without the outcome, was estimated from held-out predictions of models fitted to the full training portion of each outer fold. For conformal prediction and the longitudinal analyses, separate models were developed after 20% of the outer training portion had been set aside for calibration. Hyperparameters were selected by fivefold internal cross-validation within the remaining proper training subset, and the model was then fitted to that subset. The calibration set was used to determine the conformal thresholds. These models then produced the held-out predictions used for conformal prediction and the longitudinal analyses.

Missing continuous values were imputed using five-nearest-neighbour imputation (KNN, k = 5). Continuous predictors were scaled using the 5th and 95th percentiles of the training data, with values below the 5th percentile truncated to 0, values above the 95th percentile truncated to 1, and intermediate values linearly scaled between 0 and 1. Missing categorical values were treated as a separate category before one-hot encoding. No feature selection was performed beyond excluding variables with more than 20% missingness. Participants with the outcome formed the positive class, and those without it formed the negative class. To account for the smaller positive class, its weight was set to the square root of the negative-to-positive sample ratio in the training data; the negative-class weight was set to 1. Elastic-net hyperparameters were selected by fivefold internal cross-validation using the area under the receiver operating characteristic curve (AUROC) as the selection criterion, with l1_ratio ∈ {0.1, 0.3, 0.5, 0.7, 0.9} and C ∈ {0.001, 0.01, 0.1, 0.5, 1}. Preprocessing parameters and class weights were estimated from the training data used for each model: the full outer training portion for discrimination models and the proper training subset for conformal models. The fitted preprocessing transformations were applied unchanged to the corresponding evaluation data. Model performance was summarised by the AUROC. The area under the precision–recall curve, Brier score, and calibration intercept and slope for the predicted probabilities are reported in Supplementary Table S7.

Conformal prediction was applied separately for each outcome and antenatal visit using predicted probabilities from the corresponding visit-specific model^14^. Conformal calibration was performed separately for participants with and without the outcome, using a Mondrian split-conformal procedure^15^. For a calibration participant with outcome class *y* ∈ {0,1}, the nonconformity score was defined as *s* = 1 − *p̂_y_*, where *p̂_y_* denotes the predicted probability assigned to the observed class.

For each class y and confidence level 1 − *α*, the conformal threshold was determined from the *n_y_* calibration participants in that class using the finite-sample corrected rank *k_y_* = 01*n_y_* + 13(1 − *α*)6. The threshold *q_y_* was the *k_y_*-th smallest class-specific calibration score, with q*_y_*=+ ∞ when *k_y_* > *n_y_*. For a held-out participant, class y was included in the prediction set when 1 − *p̂_y_* ≤ *q_y_*.

For binary outcomes, the resulting prediction set could contain the negative class only {0}, both classes {0,1}, the positive class only {1}, or neither class ∅. These sets were referred to as the low-risk (L), review (R), high-risk (H) and empty (E) prediction states, respectively. For longitudinal analyses, E was grouped with R because neither gave a low- or high-risk classification; the original prediction sets were retained when evaluating coverage. Predictions that were not evaluable at a given visit were not assigned a conformal prediction state.

For the binary outcomes considered here, these states could also be expressed as thresholds on the predicted event probability *p̂*: class 0 was included when *p̂* ≤ *q*_0_, whereas class 1 was included when *p̂* ≥ 1 − *q*_1_, where *q*_0_ and *q*_1_ denote the corresponding class-specific conformal thresholds. The two thresholds therefore partitioned the predicted-probability axis into the four prediction-set states defined above. Numerical agreement between the set-based and probability-threshold representations was evaluated at each visit and for complete three-visit trajectories (Supplementary Method S2).

The primary analysis used a 90% confidence level, corresponding to a 90% coverage target, with 80% examined in a sensitivity analysis. Under exchangeability within the Mondrian partitions, the procedure targets finite-sample coverage within each outcome class at each visit^15^. Empirical coverage was evaluated at each visit overall and separately within the two outcome classes. A prediction was counted as covered when the prediction set contained the observed class; empty sets were counted as not covered.

Visits for which no evaluable conformal prediction set was produced were excluded from the visit-specific denominator. Among participants with evaluable prediction sets at all three visits, we calculated simultaneous coverage as the proportion whose observed outcome was included in the set at every visit.

### Longitudinal prediction trajectories

For each outcome, the sequence of prediction states across the three visits defined a participant’s prediction trajectory, representing her prediction history. A trajectory was included only when an evaluable state was available at all three visits.

Trajectories were grouped according to the Visit 3 state and prediction history (Table 2). Among participants in the high-risk state at Visit 3, persistent high was defined as H-H-H. Emerging high comprised trajectories that first entered the high-risk state at Visit 2 and remained high risk at Visit 3, or first entered the high-risk state at Visit 3. Intermittent high comprised trajectories that were high risk at Visit 1, moved to a review or low-risk state at Visit 2, and returned to high risk at Visit 3. Among participants in the low-risk state at Visit 3, stable low was defined as L-L-L, whereas resolved-to-low comprised trajectories with at least one previous review state but no previous high-risk state. The individual three-visit sequences within each group are reported in Supplementary Table S3.

**Table 2.** Definition of longitudinal trajectory groups based on visit-specific conformal states.

| **Visit 3 state** | **Trajectory group** | **Three-visit sequences** |
|---|---|---|
| High (H) | Persistent high | H-H-H |
| High (H) | Emerging high | L-H-H, R-H-H, L-L-H, L-R-H, R-L-H, R-R-H |
| High (H) | Intermittent high | H-L-H, H-R-H |
| Low (L) | Stable low | L-L-L |
| Low (L) | Resolved-to-low | L-R-L, R-L-L, R-R-L |
| Low (L) | Prior high to low | H-L-L, H-R-L, H-H-L, L-H-L, R-H-L |
| Review (R) | Review-ending trajectories | All evaluable sequences ending in R |

**Table note**: L, low risk; R, review; H, high risk. Each sequence represents the conformal state at Visits 1, 2, and 3, respectively.

We compared trajectory groups that shared the same Visit 3 state to examine whether the proportion with the outcome differed according to prediction history. Among participants in the high-risk state at Visit 3, the comparison was persistent high versus emerging high; among those in the low-risk state, stable low versus resolved-to-low. To account for differences in predicted probability within the same Visit 3 state, these contrasts were estimated both before and after accounting for the Visit 3 predicted probability. For each outcome, observed outcome status was modelled using logistic regression with trajectory group and the Visit 3 predicted probability, represented by a natural cubic spline with four degrees of freedom. For each contrast, adjusted risk differences were estimated by g-computation^22^, standardising predicted risks over the combined sample of the two trajectory groups being compared. Confidence intervals were obtained by participant-level bootstrap resampling.

We performed two additional analyses to compare persistent-high and emerging-high participants within a shared range of Visit 3 predicted probability. First, we restricted both groups to the overlap of their respective 5th–95th percentile ranges of Visit 3 predicted probability (common support). We then refitted the spline-adjusted model and re-estimated the risk difference in this restricted sample.

Second, we formed matched pairs containing one participant from each group using nearest-neighbour matching on the logit-transformed Visit 3 predicted probability. Matching was performed without replacement, with a caliper of 0.2 pooled standard deviations. We assessed balance in the matching variable using the standardised mean difference and calculated the risk difference from the proportions with the outcome in the matched groups. Further details are provided in Supplementary Method S3 and Supplementary Fig. S1.

### Cross-outcome associations in prediction changes

The preceding analyses followed each outcome separately. We next examined whether predictions for different outcomes changed together within the same participant, and whether such associations could be traced to predictors entering more than one model.

For each pair of outcomes, we compared how their prediction states changed between adjacent visits. States were ordered as L < R < H, and a move to a higher state was defined as an upward transition. Each outcome was classified as having an upward or non-upward transition, and the association between the two classifications was summarised using an odds ratio. Analyses were restricted to participants with evaluable states for both outcomes at both visits and were performed separately for Visit 1 to Visit 2 and Visit 2 to Visit 3. Complete pairwise upward-transition associations are reported in Supplementary Table S4.

The same question was then examined without discretising predictions into states. For each outcome we calculated the change in held-out predicted probability between adjacent visits, and summarised pairwise associations between these changes using Spearman correlation coefficients.

Where predictions for two outcomes changed together, we examined the contribution of predictors entering both models. We expressed each model’s predicted probability on the logit scale. On this scale, the model output is called the linear predictor and equals the intercept plus the sum of individual predictor contributions. A predictor was treated as shared when it entered both outcome-specific fold models for that participant at that visit. The contributions of these predictors were summed to obtain the shared component. The remaining component comprised the intercept and contributions from all other predictors, such that the shared and remaining components summed to the complete linear predictor. For the complete linear predictor and each component, we calculated the change between adjacent visits. We then correlated these changes between the two outcome models across participants using Spearman correlation coefficients.

### Statistical analysis

Categorical variables were summarised as counts and proportions. Differences between trajectory groups in the proportion with the outcome were reported as absolute risk differences (percentage points) with 95% confidence intervals. For comparisons accounting for the Visit 3 predicted probability, adjusted risk differences were estimated by g-computation. Associations between upward transitions across outcomes were summarised using odds ratios and evaluated using two-sided Fisher’s exact tests. Associations between changes in predicted probabilities or model components were summarised using Spearman correlation coefficients.

AUROC 95% confidence intervals were estimated from 2,000 participant-level bootstrap resamples of the held-out predictions using percentile intervals. Wilson intervals were used for visit-specific and three-visit simultaneous empirical coverage; the latter was evaluated among participants with evaluable prediction sets at all three visits. The three-visit independence benchmark was calculated for each outcome as the product of the three observed visit-specific marginal coverage rates among participants evaluable at all three visits. The Bonferroni lower-bound reference was calculated as 1 − 3(1 − *c*), where c was the nominal single-visit confidence level. Confidence intervals for adjusted trajectory-group risk differences and the common-support-restricted analysis used 2,000 participant-level bootstrap resamples, with held-out predicted probabilities and trajectory classifications kept fixed. The adjustment model was refitted within each resample. The matching analysis used 500 bootstrap resamples of the original matched pairs. Trajectory-analysis bootstrap intervals were defined by the 2.5th and 97.5th percentiles of the bootstrap distribution.

For pairwise cross-outcome analyses, P values were adjusted for the 10 outcome pairs within each visit interval using the Benjamini–Hochberg procedure. P values and Benjamini–Hochberg-adjusted q values are reported in Supplementary Tables S4 and S5. All analyses were performed using Python 3.12, scikit-learn 1.8.0, statsmodels 0.14.6 and SciPy 1.17.1. This study is reported in accordance with the TRIPOD+AI statement^23^. The completed checklist is provided in Supplementary Table S11. A large language model tool was used only for grammar correction, spelling checks and readability improvement. No scientific interpretations, analytical outputs, arguments or manuscript drafts were generated by the tool. All text content was fully reviewed, revised and fact-checked by the authors, who take complete responsibility for the final version of the manuscript.

## Results

### Later visits yield greater discrimination and more single-class predictions for fetal growth

Table 1 summarises the baseline characteristics of the 9,289 participants, who were enrolled at 6–13 weeks of gestation and followed to delivery. Outcome-specific analysis sets included 8,509–9,289 participants at Visits 1 and 2 and 8,453–9,262 at Visit 3 (Supplementary Table S2). Visit-specific analysis sets incorporated the outcome-specific data requirements and the Visit 3 restriction described in Methods. Outcome prevalence ranged from 2.8% for FGR (245/8,787) to 11.4% for LGA (968/8,509), with GDM, PE, and PTB occurring in 4.3% (376/8,720), 5.9% (552/9,289), and 8.7% (772/8,853), respectively.

Among participants with FGR included in the Visit 3 analysis, suspected FGR was recorded a mean of 53.1 days (SD, 23.4) after the Visit 3 assessment. Discrimination increased at each successive visit for all five outcomes, although the magnitude of improvement differed (Fig. 2a). The largest increases were observed for the fetal growth outcomes. For FGR, the AUROC increased from 0.596 (95% CI 0.560– 0.632) at Visit 1 to 0.811 (0.782–0.839) at Visit 3, while for LGA it increased from 0.629 (0.610–0.648) to 0.766 (0.750–0.781). Changes were smaller for GDM, PE, and PTB.

**Figure 2.**
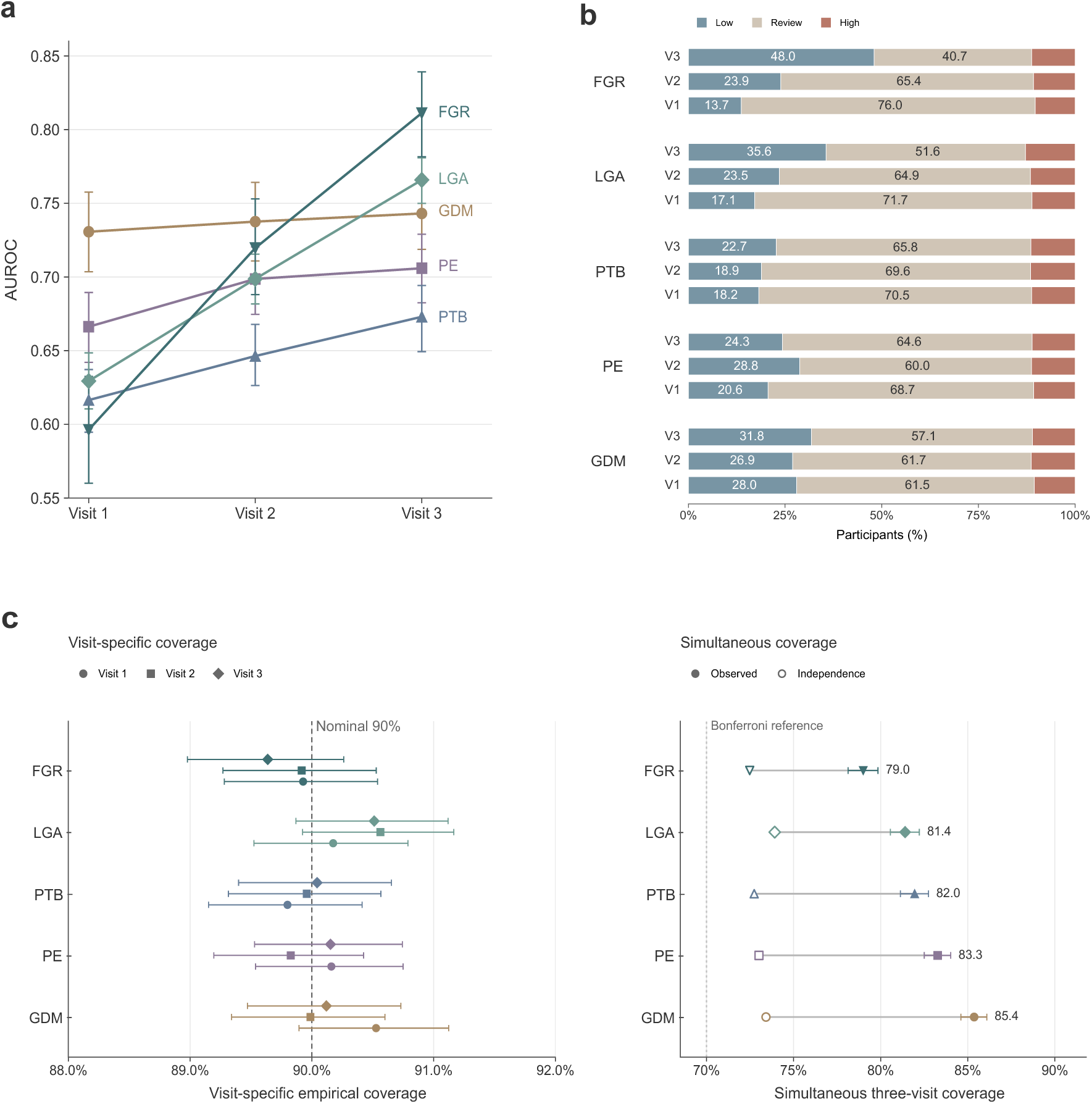
Discrimination, prediction-state composition, and empirical coverage across gestation. a, outcome-specific discrimination at Visits 1–3, summarised by the AUROC. Points show AUROC estimates and error bars indicate 95% confidence intervals. b, distribution of low-risk (L), review (R) and high-risk (H) prediction states at the 90% coverage target. Outcomes are displayed from FGR to GDM, with Visit 3 shown above Visits 2 and 1 within each outcome. Percentages are shown for the low-risk and review states. c, empirical coverage of the 90% conformal prediction sets. The left panel shows visit-specific marginal coverage, with the dashed line indicating the nominal 90% level. The right panel shows simultaneous coverage across all three visits among participants with evaluable predictions at each visit. Filled symbols indicate observed simultaneous coverage, and open symbols indicate the independence benchmark calculated as the product of the three observed visit-specific marginal coverage rates for the corresponding outcome. The dotted line indicates the 70% Bonferroni lower-bound reference. Error bars indicate 95% confidence intervals. Abbreviations: GDM, gestational diabetes mellitus; PE, pre-eclampsia; PTB, preterm birth; LGA, large for gestational age; FGR, fetal growth restriction.

Prediction-state composition also changed across gestation (Fig. 2b). At the 90% coverage target, the proportion of review predictions decreased most for FGR, from 76.0% at Visit 1 to 40.7% at Visit 3, and for LGA, from 71.7% to 51.6%. These reductions were accompanied mainly by increases in low-risk predictions, from 13.7% to 48.0% for FGR and from 17.1% to 35.6% for LGA. The proportion of high-risk predictions changed little across visits, remaining between 10.3% and 12.8% across all outcome–visit combinations. The low-risk, review and high-risk states could also be represented using two cut-offs on the predicted probability (Methods). The two representations assigned the same state to 99.9% of predictions at each visit and the same three-visit sequence to 99.7% of complete trajectories.

### Prediction-history differences after accounting for Visit 3 predicted probability

Prediction states moved both upward and downward between visits (Supplementary Table S9). Participants could reach the same prediction state at Visit 3 through different prediction histories. Among participants classified as high risk at Visit 3, we compared those classified as high risk at all three visits (persistent high) with those first classified as high risk at Visit 2 or 3 (emerging high) (Fig. 3a). Before adjustment, the proportion with the outcome was higher in the persistent-high group for GDM, PE, PTB and LGA, with risk differences of +6.8, +4.3, +5.7 and +3.1 percentage points, respectively. For FGR, the difference was −1.8 percentage points.

**Figure 3.**
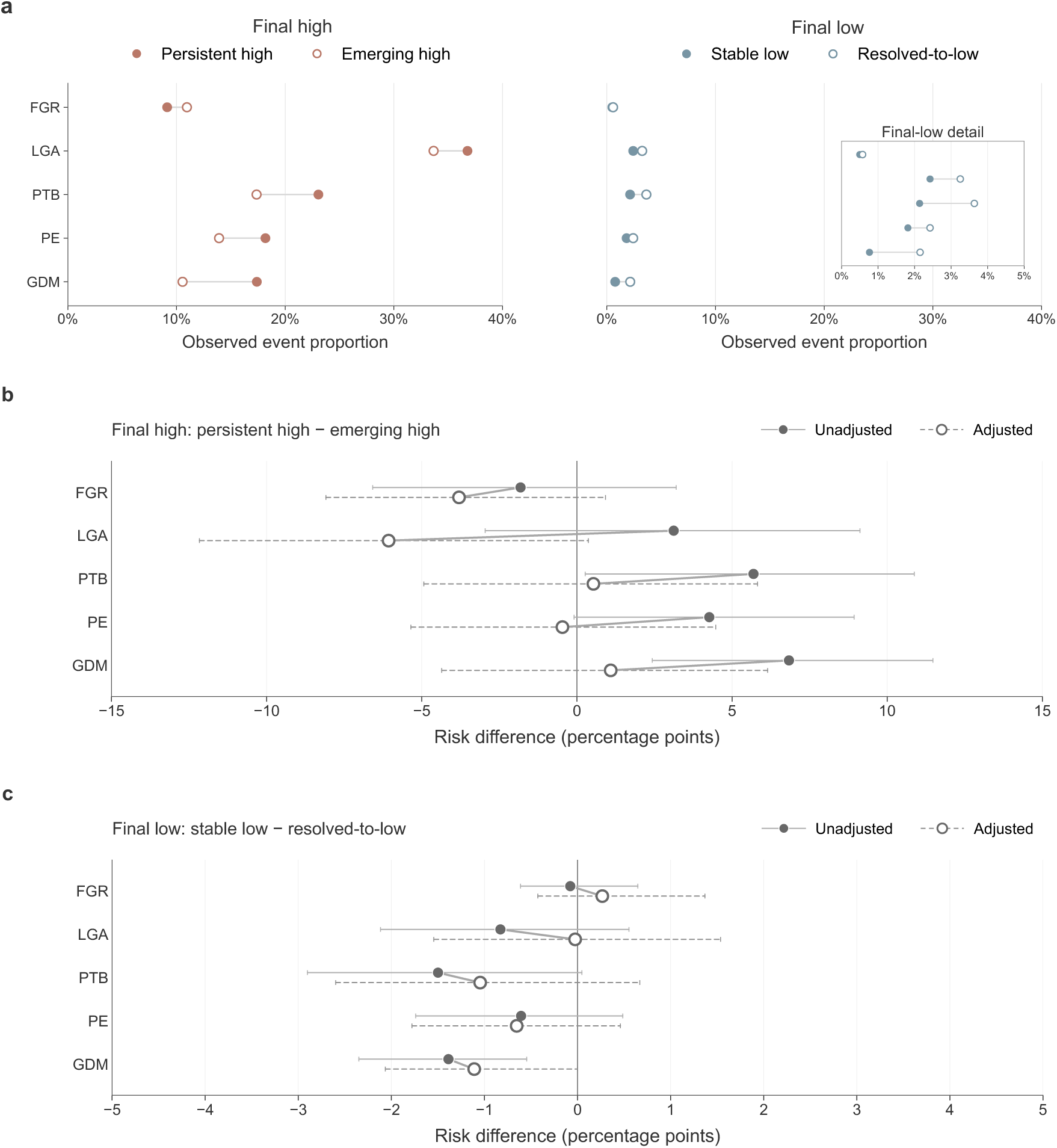
Proportion with the outcome according to longitudinal prediction history before and after accounting for Visit 3 predicted probability. a, proportion with the outcome among participants in the high-risk or low-risk state at Visit 3, stratified by their preceding prediction history. Persistent high denotes H-H-H trajectories, whereas emerging high denotes trajectories that first reached the high-risk state at Visit 2 or Visit 3. Stable low denotes L-L-L trajectories, whereas resolved-to-low denotes trajectories ending in low risk after at least one preceding review state and no preceding high-risk state. The inset shows the same final-low data on an expanded 0–5% scale. b, unadjusted and adjusted risk differences (persistent high minus emerging high) among participants in the high-risk state at Visit 3. c, corresponding risk differences (stable low minus resolved-to-low) among participants in the low-risk state at Visit 3; panel c uses an expanded x-axis (−5 to +5 percentage points) relative to panel b (−15 to +15 percentage points). Adjusted estimates account for the Visit 3 predicted probability using a natural cubic spline with four degrees of freedom. Points indicate risk differences and horizontal lines indicate 95% confidence intervals. Group sizes and constituent three-visit sequences are given in Supplementary Table S3.

Because the high-risk state spans a range of predicted probabilities, part of these differences could reflect differences in Visit 3 predicted probability between the two groups. After accounting for this probability, the estimated proportion with the outcome was higher in the emerging-high group than in the persistent-high group for LGA and FGR (Fig. 3b). Adjusted risk differences, calculated as persistent-high minus emerging-high, were −6.1 percentage points (95% CI −12.2 to 0.4) for LGA and −3.8 (−8.1 to 0.9) for FGR. The LGA estimate was similar after common-support restriction and direct matching (Supplementary Fig. S1). For GDM, PE and PTB, the corresponding adjusted differences were +1.1 percentage points (95% CI −4.4 to 6.2), −0.5 (−5.4 to 4.5) and +0.5 (−4.9 to 5.8), respectively. Estimates at the 80% coverage target and the corresponding trajectory-group composition are reported in Supplementary Tables S8 and S9.

Participants who delivered before 30+0 weeks had no Visit 3 prediction and were therefore not included in these comparisons: 95 in the FGR analysis and 56 in the LGA analysis, of whom 9 and 6, respectively, had the outcome. At the 90% coverage target, 6 of these participants in the FGR analysis and 7 in the LGA analysis had high-risk predictions at both Visit 1 and Visit 2, and 2 in each group had the corresponding outcome.

Among participants classified as low risk at Visit 3, we compared those classified as low risk at all three visits (stable low) with those who had at least one earlier review prediction and no earlier high-risk prediction (resolved-to-low). The proportion with the outcome ranged from 0.5% to 3.6% across both groups and all five outcomes (Fig. 3a). After accounting for Visit 3 predicted probability, the differences (stable low minus resolved-to-low) ranged from −1.1 to +0.3 percentage points (Fig. 3c).

### Shared predictor contributions help explain linked changes across outcomes

The preceding analyses considered each outcome separately, although predictions for several outcomes were generated for the same participants at each visit. We therefore examined whether prediction changes between consecutive visits were associated across outcomes. We first considered whether a participant’s prediction state moved to a higher level, from low risk to review or high risk, or from review to high risk (Fig. 4a). Between Visits 1 and 2, upward changes in PTB and FGR tended to occur together (OR, 3.26), whereas upward changes in LGA were less likely to coincide with those in FGR (OR, 0.08) or PTB (OR, 0.27). Between Visits 2 and 3, the association remained positive for PTB and FGR (OR, 1.88) and negative for LGA and FGR (OR, 0.05; Supplementary Table S4). No participant had both LGA and FGR.

**Figure 4.**
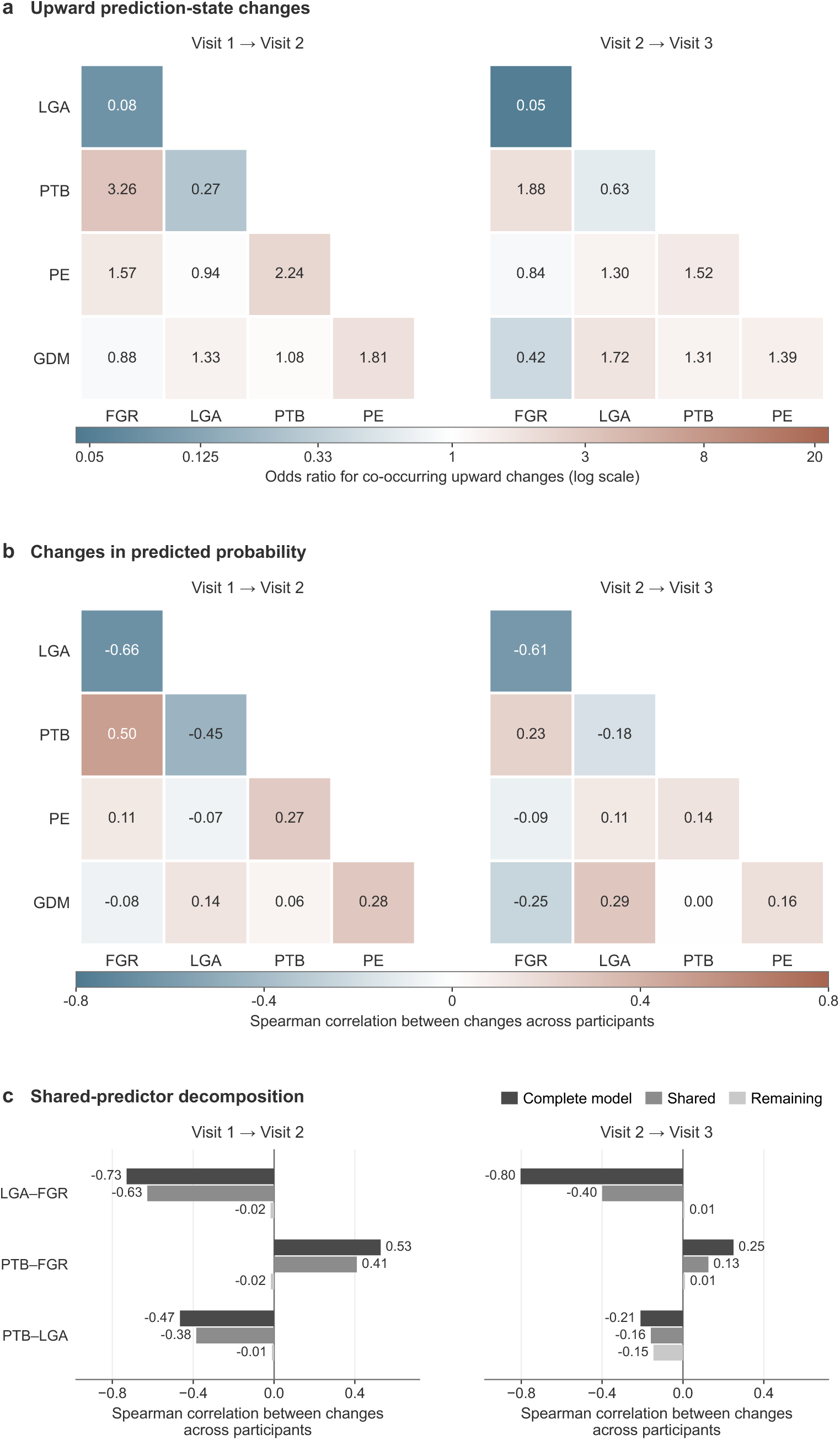
Associations between prediction changes across outcomes and the contribution of shared predictors. a, association between upward changes in prediction state for each pair of outcomes, between Visit 1 and Visit 2 (left) and between Visit 2 and Visit 3 (right). An upward change denotes a move to a higher state in the ordering low risk < review < high risk. Cells show odds ratios for the association between upward transitions in the two outcomes. The colour scale is symmetric on the logarithmic scale. b, Spearman correlations between changes in predicted probability for each pair of outcomes, calculated across participants over the same two intervals. c, Spearman correlations between changes in the outputs of paired outcome-specific models, calculated separately for the complete linear predictor, the shared-predictor component and the remaining component. All three components were evaluated on the logit scale. The three outcome pairs with the strongest associations in panels a and b are shown; bars extend from zero, with correlation coefficients shown at their ends. State-transition analyses in panel a used the 90% coverage target. All panels used held-out predictions and included participants with evaluable predictions for both outcomes at both visits. Complete state-transition associations, probability-change correlations and shared-predictor decompositions are reported in Supplementary Tables S4–S6, respectively. Abbreviations: GDM, gestational diabetes mellitus; PE, pre-eclampsia; PTB, preterm birth; LGA, large for gestational age; FGR, fetal growth restriction.

For each participant and outcome, we next calculated the change in predicted probability between consecutive visits and examined whether these changes were correlated between pairs of outcomes across participants (Fig. 4b). Between Visits 1 and 2, changes in predicted PTB and FGR risk were positively correlated (Spearman ρ = 0.50): participants whose predicted PTB risk increased tended also to have an increase in predicted FGR risk. Changes in predicted LGA and FGR risk were negatively correlated (ρ = −0.66), as were changes in PTB and LGA (ρ = −0.45). Between Visits 2 and 3, the corresponding correlations were 0.23, −0.61 and −0.18, respectively. The association between LGA and FGR was similar across both intervals, whereas those involving PTB were weaker later in gestation (Supplementary Table S5).

We next examined which model components contributed to these cross-outcome patterns. On the logit scale, each model’s linear predictor is the sum of an intercept and individual predictor contributions. For each outcome pair, we separated contributions from predictors shared by both models from a remainder containing the intercept and other predictors, then correlated changes in each component between the two outcome models (Fig. 4c; Supplementary Table S6). For LGA and FGR between Visits 1 and 2, changes in the complete linear predictor were correlated at −0.73, compared with −0.63 for the shared-predictor component and −0.02 for the remaining component. A similar pattern was observed for PTB and FGR (0.53 overall, 0.41 for the shared component and −0.02 for the remainder). For these pairs, changes in the shared-predictor component were correlated in the same direction and with a similar pattern as the complete linear predictor, whereas correlations for the remaining component were close to zero. Because a shared predictor could enter two models with coefficients of opposite sign, shared information could contribute to either concordant or discordant prediction changes.

### Coverage is close to target at each visit and lower across all three visits

Visit-specific empirical coverage remained close to the nominal 90% level across all outcomes and visits, ranging from 89.6% to 90.6% (Fig. 2c). At the 90% coverage target, observed coverage ranged from 89.7% to 90.5% among participants without the outcome and from 87.7% to 92.8% among those with the outcome across outcomes and visits (Supplementary Table S10). Among participants with evaluable prediction sets at all three visits, simultaneous coverage ranged from 79.0% for FGR to 85.4% for GDM, below the 90% per-visit target for every outcome. In each case, it exceeded both the corresponding independence benchmark, calculated from the product of the three observed marginal coverage rates among participants evaluable at all three visits (72.5–73.9%), and the 70% Bonferroni lower-bound reference implied by three 90% visit-specific targets. At least one prediction set excluded the actual outcome in 15–21% of participants, compared with 26–28% expected under independence. These failures tended to recur in the same participants, rather than occur independently across visits. The excess above the independence benchmark was largest for GDM (+12.0 percentage points), which had the smallest increase in AUROC from Visit 1 to Visit 3, and smallest for FGR (+6.5 percentage points), which had the largest increase.

## Discussion

Among women classified as high risk at Visit 3, we compared those first classified as high risk at Visit 2 or 3 (emerging high) with those classified as high risk at all three visits (persistent high). After accounting for Visit 3 predicted probability, the estimated proportion with the outcome was higher in the emerging-high group for both LGA and FGR. The estimated differences were 6.1 percentage points (95% CI −0.4 to 12.2) for LGA and 3.8 (−0.9 to 8.1) for FGR. For GDM, PE and PTB, the adjusted point estimates were close to zero. Changes in predictions were also associated across outcomes, with the strongest associations reflected mainly in shared predictor contributions. Prediction sets included the actual outcome in close to 90% of women at each visit, as targeted. Among women with predictions at all three visits, the set included the actual outcome at every visit in 79.0–85.4%, depending on the outcome.

At the 90% coverage target, conformal prediction separated low- and high-risk predictions from review predictions. A review state meant that conformal prediction did not give a low- or high-risk classification at the chosen coverage target. The boundaries of this state were set by conformal calibration rather than by clinical thresholds for intermediate risk. In our study, the gains in discrimination for FGR and LGA coincided with fewer review predictions and more low-risk predictions, while empirical coverage remained close to the target. For antenatal assessment, the practical question is how to present this review state alongside other clinical information and whether and when it should prompt reassessment.

Discrimination improved most for FGR and LGA as measurements accumulated across visits, and less for GDM, PE and PTB. Studies of individual markers help put the smaller PTB and PE gains in context. In nuMoM2b, serial cervical length and fetal fibronectin measurements had limited predictive accuracy for spontaneous PTB^24^. Uterine artery Doppler measurements were associated with PE but had limited predictive performance in a separate low-risk nulliparous cohort^25^.

In our study, women classified the same way at Visit 3 differed in their earlier predictions. For fetal growth, the adjusted comparisons suggest that prediction history may help interpret the Visit 3 predicted probability. The LGA estimate changed little after restricting the comparison to the shared probability range or matching on Visit 3 predicted probability. The two fetal growth outcomes also differed in ascertainment: LGA was determined from birthweight, whereas FGR reflected a clinical suspicion recorded during pregnancy, so the FGR comparison also depends on when that suspicion was recorded. For GDM, PE and PTB, adjustment for Visit 3 predicted probability brought the estimated history-group differences close to zero. Adjusted differences between stable-low and resolved-to-low histories were also close to zero. These findings raise the question of what information distinguished emerging-high from persistent-high histories for fetal growth.

For fetal growth, change over time may provide information beyond current size^26^. In the POP study, the association between low estimated fetal weight and neonatal morbidity varied with abdominal circumference growth velocity^27^. In our study, similar Visit 3 predicted probabilities may have arisen from different combinations of early maternal characteristics and later fetal measurements. Earlier predictions may help distinguish these combinations. A next step would be to test whether explicitly modelling fetal growth over time changes the adjusted history-group differences.

Information shared across models also helped explain how predictions for different outcomes changed together. For the most strongly associated outcome pairs, correlations between changes in shared predictor contributions had the same direction as correlations between changes in the complete linear predictor, whereas correlations between changes in the remaining contributions were close to zero. A shared predictor’s contribution depends on both its value and its coefficient in each visit-specific model. Shared predictors can therefore contribute to changes in the same or opposite directions across outcomes, as observed for PTB and FGR, and for LGA and FGR. Presenting related predictions alongside their shared predictor contributions could help clinicians understand why several risk estimates change together.

Across the complete visit sequence, 15–21% of women with prediction sets at all three visits had at least one visit at which the set did not include the actual outcome, compared with 26–28% expected if coverage at the three visits had been independent. Conformal calibration at each visit targets the prespecified coverage for that visit; coverage across all three visits also depends on whether prediction sets exclude the actual outcome repeatedly in the same women. Repeated use of earlier measurements in later models may help explain why the same women were affected at several visits. Conformal methods developed for time-series prediction address related coverage problems^28^. Temporal Quantile Adjustment addresses cross-sectional and longitudinal coverage in time-series regression^29^. CopulaCPTS addresses coverage for forecasts of multiple future responses from a common origin, using dependence among forecast errors^30^. In our study, successive models predicted the same final binary outcome as new information became available. The next methodological question is how to target coverage across this visit sequence while keeping prediction sets informative.

The clinical role of a prediction depends on when it is made. Early preventive interventions, as in the ASPRE trial, require early risk identification^31^; later predictions may inform surveillance and assessment. The handling of a review state would therefore need to reflect the decisions available at that visit. For fetal growth outcomes, a high-risk prediction at Visit 3 could be displayed alongside earlier predictions, including the visit at which the woman was first classified as high risk. Clinical evaluation could then examine whether these histories should influence the choice or timing of reassessment. Once a complication has been diagnosed, predicting its first occurrence is no longer the relevant task, although other complications may still require risk assessment.

Before these prediction states are used in another population, their coverage and the proportions of low-risk, review and high-risk predictions would need to be evaluated there. In an external validation of a conformal sepsis model, discrimination declined and prediction sets containing more than one possible answer became more frequent^32^. Independent, labelled calibration data representative of the local population can support recalibration to a locally specified coverage target.

When several outcomes are assessed together, their predictions may lead to one shared assessment or to separate clinical actions. Changes in the same measurements can raise two risk estimates that call for different interventions. If predictions are used to generate flags, the number of flags, the number of women flagged and the number of resulting clinical actions describe different aspects of workload. In a study of clinical decision support, more alerts, particularly repeated alerts, were associated with lower acceptance rates^33^. Combining predictions into clinical actions would therefore require considering outcome priorities, the timing of available actions and local resources.

The study used one cohort of nulliparous women with singleton pregnancies and had no independent external validation. Model performance, history-group differences and coverage remain to be evaluated in other populations, and the history comparisons may also depend on the predictors and model structure used. Outcome definitions and timing also affect interpretation. For all FGR cases included at Visit 3, suspicion was recorded after the Visit 3 assessment; because the database did not record the examinations on which each suspicion was based, the extent of any overlap with information already available at Visit 3 could not be determined. Routine GDM diagnosis overlapped Visit 3, and some PE diagnoses may also have been made by that visit. Because deliveries before 30+0 weeks were excluded, Visit 3 results, including PTB results and comparisons of prediction histories, apply to pregnancies continuing to at least that gestational age. We did not evaluate how using these predictions changed clinical decisions, resource use or patient outcomes.

Among women classified as high risk at Visit 3, the adjusted proportion with the outcome was higher in the emerging-high group than in the persistent-high group for both LGA and FGR. The contribution analysis traced linked changes across outcomes to shared model inputs. The coverage results show that repeated predictions need to be evaluated across the complete visit sequence as well as at each visit. This approach can inform other longitudinal, multi-outcome classification studies, with contribution analysis adapted to the model structure. Clinical evaluation can then address whether and when review predictions should lead to reassessment and how related predictions are combined into decisions.

## Data availability

Data from the nuMoM2b study are available from the NICHD Data and Specimen Hub (DASH) at https://dash.nichd.nih.gov, subject to approval by the DASH Data Access Committee. Variable definitions and case report forms are distributed with the dataset.

## Code availability

Code for model development, prediction, conformal calibration and the downstream analyses reported in this study is available at https://github.com/yanqi-tue/conformal_prediction_yq. The code operates on processed analytical datasets derived from the controlled-access nuMoM2b data. Neither the original data nor the processed analytical datasets are included in the repository or permitted for redistribution. Access to the data required to run the code must be obtained independently through NICHD DASH, as described in the Data availability statement.

## Contributors

XL and YW conceived the study. YW, SA, and XL designed the analysis, wrote the code, and did the statistical analyses. YW, XL, and QL interpreted the results. MBvdH, SGO, MX, JOEHvL, and CC provided clinical interpretation. YW wrote the first draft of the manuscript. EP, MvdV, MBvdH, MX, QL, CC, JOEHvL, SGO, and MM reviewed and edited the manuscript. XL and MM supervised the study. YW and XL accessed and verified the underlying data. All authors had full access to all the data in the study, read and approved the final manuscript, and accepted responsibility to submit for publication.

## Acknowledgements

This study received no specific funding. This study was performed within the framework of the Eindhoven MedTech Innovation Center (e/MTIC). e/MTIC had no role in study design, data collection, data analysis, data interpretation, or manuscript preparation.

