## Supplemental Materials for "Uncertainty-aware longitudinal prediction of multiple adverse pregnancy outcomes"

### Supplementary Methods

#### Supplementary Method S1. Gestational diabetes mellitus diagnostic criteria

GDM was defined by one of the following glucose tolerance test criteria: a 3-h 100-g oral glucose tolerance test with at least two values meeting or exceeding 95 mg/dL fasting, 180 mg/dL at 1 h, 155 mg/dL at 2 h or 140 mg/dL at 3 h; a 2-h 75-g oral glucose tolerance test with at least one value meeting or exceeding 92 mg/dL fasting, 180 mg/dL at 1 h or 153 mg/dL at 2 h; or, when neither of these fasting tests was performed, a non-fasting 50-g glucose challenge result of at least 200 mg/dL. When no glucose tolerance test results were available, GDM recorded in the medical chart was used. Participants with pregestational diabetes were excluded from GDM analyses. These criteria follow the nuMoM2b study definitions (Facco et al., Obstet. Gynecol. 2017; main-text reference 20).

#### Supplementary Method S2. Probability-threshold representation and numerical verification

For a binary outcome, let p denote the predicted probability of the event class. With nonconformity score equal to 1 minus the probability assigned to the observed class, class 0 was included in the conformal prediction set when p was no greater than the class-0 calibration threshold, whereas class 1 was included when 1−p was no greater than the class-1 calibration threshold. The resulting prediction set could therefore be represented as a partition of the predicted-probability axis using two class-specific thresholds. The singleton set containing class 0 was labeled low (L), the set containing both classes review (R), the singleton set containing class 1 high (H), and the empty set empty (E). For longitudinal analyses, E was grouped with R because neither state provided a singleton prediction; the original prediction sets were retained for coverage calculations. Numerical equivalence of the set-based and probability-threshold representations was evaluated by comparing participant-level states at each visit and the resulting complete three-visit trajectories. At the primary 90% confidence level, agreement was 99.9% at each visit and 99.7% for complete three-visit trajectories; the small discrepancies were confined to numerical threshold-edge cases.

#### Supplementary Method S3. Sensitivity analyses for prediction-history comparisons

Common-support restriction used the intersection of the 5th–95th percentile ranges of Visit 3 predicted probability in the persistent-high and emerging-high groups. Matching was 1:1 greedy nearest-neighbor matching without replacement on the logit of Visit 3 predicted probability, with a caliper of 0.2 pooled standard deviations of this logit. Balance was assessed by the standardized mean difference in this logit before and after matching. Confidence intervals for the matched risk difference were obtained from 500 bootstrap resamples of matched pairs.

**Supplementary Tables**

**Table S1. Candidate predictors included at each antenatal visit**

| Predictor domain | Candidate predictors | Visit 1 (6–13 weeks) | Visit 2 (16–21 weeks) | Visit 3 (22–29 weeks) |
|---|---|---|---|---|
| **Demographic characteristics** | Maternal age; race/ethnicity; income level; education level | ✓ | ↳ | ↳ |
| **Baseline anthropometry** | Pre-pregnancy BMI; waist circumference; waist circumference over the iliac crest; hip circumference; neck circumference | ✓ | ↳ | ↳ |
| **Maternal weight** | Maternal weight | — | ✓ | ✓ |
| **Blood pressure** | Systolic and diastolic blood pressure | ✓ | ✓ | ✓ |
| **Medical and family history** | Polycystic ovary syndrome; family history of diabetes | ✓ | ↳ | ↳ |
| **Mental health** | Edinburgh Postnatal Depression Scale (EPDS) score; history of depression, anxiety, bipolar disorder, post-traumatic stress disorder, and schizophrenia/schizoaffective disorder before pregnancy | ✓ | ↳ | ✓[†] |
| **Lifestyle and sleep** | Smoking in the 3 months before the visit; snoring at or before Visit 1 | ✓ | ✓ | ✓ |
| **Diet quality** | Healthy Eating Index (HEI) components and total score; Alternative Healthy Eating Index (AHEI) components and total score | ✓ | ↳ | ↳ |
| **Pregnancy symptoms/events** | Vaginal bleeding | ✓ | ✓ | ✓ |
| **Ultrasound timing** | Gestational age at ultrasound examination | — | ✓ | ✓ |
| **Fetal biometry** | Biparietal diameter (BPD); head circumference (HC); abdominal circumference (AC); femur length (FL); estimated fetal weight (EFW)[‡] | — | ✓ | ✓ |
| **Cervical assessment** | Cervical length | — | ✓ | ✓ |

| Predictor domain | Candidate predictors | Visit 1 (6–13 weeks) | Visit 2 (16–21 weeks) | Visit 3 (22–29 weeks) |
|---|---|---|---|---|
| **Uterine artery Doppler** | Bilateral uterine artery heart rate, angle of insonation, peak systolic velocity, systolic/diastolic ratio, resistance index, pulsatility index, systolic acceleration time, and depth of diastolic notch | — | ✓ | ✓ |
| **Amniotic fluid** | Amniotic fluid measurements in four quadrants | — | — | ✓ |

**Abbreviations:** AHEI, Alternative Healthy Eating Index; BMI, body mass index; BPD, biparietal diameter; EFW, estimated fetal weight; EPDS, Edinburgh Postnatal Depression Scale; FL, femur length; HC, head circumference; HEI, Healthy Eating Index.

**Symbols:** ✓, measured or ascertained by the corresponding visit and included in the predictor set; ↳, information obtained at an earlier visit and retained in the predictor set; —, not available by the corresponding visit.

† The Visit 3 predictor set included the Visit 3 EPDS score, while baseline mental-health history was retained from Visit 1.
‡ EFW was calculated from ultrasound biometry using the Hadlock formula.

**Supplementary Table S2. Outcome-specific analysis sets and exclusion criteria**

| Outcome | Visits 1–2, n | Outcome ascertainment requirement | Exclusions | Visit 3, n |
|---|---|---|---|---|
| GDM | 8,720 | Evaluable GDM status | Missing GDM status; pre-existing diabetes | 8,623 |
| PE | 9,289 | PE status | None | 9,262 |
| PTB | 8,853 | Gestational age at delivery | Missing or invalid gestational age at delivery | 8,748 |
| LGA | 8,509 | Gestational age at delivery, birthweight and fetal sex | Missing variables or ineligibility for the birthweight reference | 8,453 |
| FGR | 8,787 | nuMoM2b FGR label | Missing FGR label; at Visit 3, suspected FGR recorded on or before the Visit 3 assessment (n = 28) | 8,666 |

Visit 3 analyses were restricted to pregnancies with delivery at or after 30+0 weeks.

**Supplementary Table S3. Longitudinal trajectory groups, constituent sequences, and group sizes**

Values are n (% of participants with an evaluable three-visit trajectory for the corresponding outcome).

| Trajectory group | Constituent sequences | GDM | PE | PTB | LGA | FGR |
|---|---|---|---|---|---|---|
| Persistent high | H-H-H | 552 (6.4%) | 462 (5.0%) | 386 (4.4%) | 310 (3.7%) | 186 (2.1%) |
| Emerging high | L-H-H; R-H-H; L-L-H; L-R-H; R-L-H; R-R-H | 341 (4.0%) | 503 (5.4%) | 547 (6.3%) | 722 (8.5%) | 721 (8.3%) |
| Intermittent high | H-L-H; H-R-H | 60 (0.7%) | 64 (0.7%) | 68 (0.8%) | 52 (0.6%) | 67 (0.8%) |
| Stable low | L-L-L | 1,440 (16.7%) | 1,046 (11.3%) | 655 (7.5%) | 743 (8.8%) | 603 (7.0%) |
| Resolved-to-low | L-R-L; R-L-L; R-R-L | 1,302 (15.1%) | 1,197 (12.9%) | 1,320 (15.1%) | 2,184 (25.8%) | 3,301 (38.1%) |
| Prior high to low | H-L-L; H-R-L; H-H-L; L-H-L; R-H-L | 2 (0.0%) | 7 (0.1%) | 12 (0.1%) | 83 (1.0%) | 257 (3.0%) |
| Review-ending | All evaluable sequences ending in R | 4,926 (57.1%) | 5,983 (64.6%) | 5,760 (65.8%) | 4,359 (51.6%) | 3,531 (40.7%) |

Complete-trajectory denominators were 8,623 for GDM, 9,262 for PE, 8,748 for PTB, 8,453 for LGA and 8,666 for FGR. L, low; R, review; H, high.

**Supplementary Table S4. Pairwise associations between upward prediction-state transitions**

An upward transition denotes movement to a higher state in the ordering $L < R < H$. P values are from two-sided Fisher exact tests; q values use Benjamini–Hochberg adjustment across the 10 outcome pairs within each interval.

| Interval | Outcome pair | N | Odds ratio | Fisher P | BH q |
|---|---|---|---|---|---|
| V1→V2 | GDM–PE | 8,720 | 1.81 | $7.3 \times 10^{-9}$ | $1.5 \times 10^{-8}$ |
| V1→V2 | GDM–PTB | 8,669 | 1.08 | 0.46 | 0.51 |
| V1→V2 | GDM–LGA | 8,375 | 1.33 | 0.0052 | 0.0074 |
| V1→V2 | GDM–FGR | 8,649 | 0.88 | 0.27 | 0.34 |
| V1→V2 | PE–PTB | 8,853 | 2.24 | $4.4 \times 10^{-16}$ | $1.1 \times 10^{-15}$ |
| V1→V2 | PE–LGA | 8,509 | 0.94 | 0.67 | 0.67 |
| V1→V2 | PE–FGR | 8,787 | 1.57 | $2.1 \times 10^{-5}$ | $3.4 \times 10^{-5}$ |
| V1→V2 | PTB–LGA | 8,509 | 0.27 | $1.5 \times 10^{-18}$ | $4.9 \times 10^{-18}$ |
| V1→V2 | PTB–FGR | 8,777 | 3.26 | $8.5 \times 10^{-42}$ | $8.5 \times 10^{-41}$ |
| V1→V2 | LGA–FGR | 8,502 | 0.08 | $4.0 \times 10^{-37}$ | $2.0 \times 10^{-36}$ |
| V2→V3 | GDM–PE | 8,613 | 1.39 | 0.0014 | 0.0021 |
| V2→V3 | GDM–PTB | 8,577 | 1.31 | 0.018 | 0.021 |
| V2→V3 | GDM–LGA | 8,323 | 1.72 | $1.0 \times 10^{-6}$ | $2.1 \times 10^{-6}$ |
| V2→V3 | GDM–FGR | 8,534 | 0.42 | $6.4 \times 10^{-8}$ | $2.1 \times 10^{-7}$ |
| V2→V3 | PE–PTB | 8,737 | 1.52 | $7.7 \times 10^{-7}$ | $1.9 \times 10^{-6}$ |
| V2→V3 | PE–LGA | 8,442 | 1.30 | 0.0029 | 0.0037 |
| V2→V3 | PE–FGR | 8,656 | 0.84 | 0.11 | 0.11 |
| V2→V3 | PTB–LGA | 8,453 | 0.63 | $3.0 \times 10^{-5}$ | $5.0 \times 10^{-5}$ |

| Interval | Outcome pair | N | Odds ratio | Fisher P | BH q |
|---|---|---|---|---|---|
| V2→V3 | PTB–FGR | 8,656 | 1.88 | $2.6 \times 10^{-11}$ | $1.3 \times 10^{-10}$ |
| V2→V3 | LGA–FGR | 8,423 | 0.05 | $1.1 \times 10^{-43}$ | $1.1 \times 10^{-42}$ |

**Supplementary Table S5. Pairwise associations between changes in predicted probability**

Correlations were calculated across participants using the change in held-out predicted probability between adjacent visits. q values use Benjamini–Hochberg adjustment across the 10 outcome pairs within each interval.

| Interval | Outcome pair | N | Spearman ρ | Spearman P | BH q |
|---|---|---|---|---|---|
| V1→V2 | GDM–PE | 8,720 | 0.282 | $1.5 \times 10^{-159}$ | $3.8 \times 10^{-159}$ |
| V1→V2 | GDM–PTB | 8,669 | 0.056 | $1.6 \times 10^{-7}$ | $1.6 \times 10^{-7}$ |
| V1→V2 | GDM–LGA | 8,375 | 0.141 | $1.9 \times 10^{-38}$ | $3.1 \times 10^{-38}$ |
| V1→V2 | GDM–FGR | 8,649 | −0.080 | $6.6 \times 10^{-14}$ | $8.2 \times 10^{-14}$ |
| V1→V2 | PE–PTB | 8,853 | 0.270 | $4.1 \times 10^{-148}$ | $8.2 \times 10^{-148}$ |
| V1→V2 | PE–LGA | 8,509 | −0.074 | $9.2 \times 10^{-12}$ | $1.0 \times 10^{-11}$ |
| V1→V2 | PE–FGR | 8,787 | 0.115 | $4.5 \times 10^{-27}$ | $6.4 \times 10^{-27}$ |
| V1→V2 | PTB–LGA | 8,509 | −0.448 | $<1 \times 10^{-300}$ | $<1 \times 10^{-300}$ |
| V1→V2 | PTB–FGR | 8,777 | 0.497 | $<1 \times 10^{-300}$ | $<1 \times 10^{-300}$ |
| V1→V2 | LGA–FGR | 8,502 | −0.662 | $<1 \times 10^{-300}$ | $<1 \times 10^{-300}$ |
| V2→V3 | GDM–PE | 8,613 | 0.163 | $1.4 \times 10^{-52}$ | $2.4 \times 10^{-52}$ |
| V2→V3 | GDM–PTB | 8,577 | 0.003 | 0.76 | 0.76 |
| V2→V3 | GDM–LGA | 8,323 | 0.292 | $8.4 \times 10^{-164}$ | $4.2 \times 10^{-163}$ |
| V2→V3 | GDM–FGR | 8,534 | −0.254 | $1.8 \times 10^{-125}$ | $6.1 \times 10^{-125}$ |
| V2→V3 | PE–PTB | 8,737 | 0.144 | $7.7 \times 10^{-42}$ | $1.1 \times 10^{-41}$ |
| V2→V3 | PE–LGA | 8,442 | 0.114 | $7.4 \times 10^{-26}$ | $9.2 \times 10^{-26}$ |
| V2→V3 | PE–FGR | 8,656 | −0.093 | $4.9 \times 10^{-18}$ | $5.4 \times 10^{-18}$ |
| V2→V3 | PTB–LGA | 8,453 | −0.182 | $4.5 \times 10^{-64}$ | $8.9 \times 10^{-64}$ |
| V2→V3 | PTB–FGR | 8,656 | 0.233 | $6.1 \times 10^{-107}$ | $1.5 \times 10^{-106}$ |
| V2→V3 | LGA–FGR | 8,423 | −0.613 | $<1 \times 10^{-300}$ | $<1 \times 10^{-300}$ |

**Supplementary Table S6. Shared-predictor decomposition of cross-outcome changes**

Values are Spearman correlations across participants between changes in the corresponding logit-scale components: the complete linear predictor, the shared-predictor component and the remaining component. The remaining component contains the intercept and contributions of predictors not shared by the two outcome models. The median number of shared predictors is reported at the start and end visit of each interval.

| Interval | Outcome pair | N | Shared predictors, start | Shared predictors, end | Complete linear predictor ρ | Shared ρ | Remaining component ρ |
|---|---|---|---|---|---|---|---|
| V1→V2 | GDM–PE | 8,720 | 54 | 83 | 0.307 | 0.234 | 0.021 |
| V2→V3 | GDM–PE | 8,613 | 83 | 101 | 0.182 | 0.077 | −0.010 |
| V1→V2 | GDM–PTB | 8,669 | 54 | 83 | 0.065 | 0.074 | −0.019 |
| V2→V3 | GDM–PTB | 8,577 | 83 | 101 | 0.003 | −0.011 | −0.015 |

| **Interval** | **Outcome pair** | **N** | **Shared predictors, start** | **Shared predictors, end** | **Complete linear predictor ρ** | **Shared ρ** | **Remaining component ρ** |
|---|---|---|---|---|---|---|---|
| V1→V2 | GDM–LGA | 8,375 | 54 | 83 | 0.142 | 0.128 | −0.002 |
| V2→V3 | GDM–LGA | 8,323 | 83 | 101 | 0.307 | 0.036 | −0.164 |
| V1→V2 | GDM–FGR | 8,649 | 54 | 83 | −0.082 | −0.056 | 0.016 |
| V2→V3 | GDM–FGR | 8,534 | 83 | 101 | −0.320 | −0.132 | 0.023 |
| V1→V2 | PE–PTB | 8,853 | 57 | 83 | 0.271 | 0.164 | −0.005 |
| V2→V3 | PE–PTB | 8,737 | 83 | 101 | 0.154 | 0.095 | 0.040 |
| V1→V2 | PE–LGA | 8,509 | 54 | 83 | −0.081 | −0.070 | 0.002 |
| V2→V3 | PE–LGA | 8,442 | 83 | 101 | 0.121 | 0.080 | 0.015 |
| V1→V2 | PE–FGR | 8,787 | 54 | 83 | 0.124 | 0.090 | 0.002 |
| V2→V3 | PE–FGR | 8,656 | 83 | 101 | −0.150 | −0.087 | 0.004 |
| V1→V2 | PTB–LGA | 8,509 | 54 | 83 | −0.466 | −0.385 | −0.010 |
| V2→V3 | PTB–LGA | 8,453 | 83 | 101 | −0.210 | −0.159 | −0.146 |
| V1→V2 | PTB–FGR | 8,777 | 54 | 83 | 0.527 | 0.409 | −0.016 |
| V2→V3 | PTB–FGR | 8,656 | 83 | 101 | 0.250 | 0.126 | 0.010 |
| V1→V2 | LGA–FGR | 8,502 | 54 | 84 | −0.729 | −0.627 | −0.017 |
| V2→V3 | LGA–FGR | 8,423 | 84 | 101 | −0.803 | −0.400 | 0.007 |

**Supplementary Table S7. Additional outcome-specific model-performance and calibration metrics**

| Outcome | Visit | AUROC (95% CI) | AUPRC | Brier score | Calibration intercept | Calibration slope |
|---|---|---|---|---|---|---|
| GDM | V1 | 0.731 (0.704–0.758) | 0.124 | 0.059 | −1.543 | 0.969 |
| GDM | V2 | 0.738 (0.711–0.764) | 0.134 | 0.059 | −1.536 | 0.925 |
| GDM | V3 | 0.743 (0.719–0.768) | 0.147 | 0.058 | −1.530 | 0.922 |
| PE | V1 | 0.666 (0.642–0.689) | 0.128 | 0.075 | −1.375 | 0.906 |

| | | | | | | |
|---|---|---|---|---|---|---|
| PE | V2 | 0.699 (0.675–0.720) | 0.155 | 0.075 | −1.361 | 0.909 |
| PE | V3 | 0.706 (0.682–0.729) | 0.154 | 0.072 | −1.397 | 0.892 |
| PTB | V1 | 0.616 (0.595–0.637) | 0.149 | 0.101 | −1.170 | 0.870 |
| PTB | V2 | 0.646 (0.626–0.668) | 0.165 | 0.098 | −1.187 | 0.860 |
| PTB | V3 | 0.673 (0.649–0.694) | 0.178 | 0.089 | −1.232 | 0.878 |
| LGA | V1 | 0.629 (0.610–0.648) | 0.174 | 0.121 | −1.024 | 0.891 |
| LGA | V2 | 0.699 (0.682–0.715) | 0.233 | 0.116 | −1.012 | 1.049 |
| LGA | V3 | 0.766 (0.750–0.781) | 0.333 | 0.107 | −1.008 | 1.043 |
| FGR | V1 | 0.596 (0.560–0.632) | 0.041 | 0.043 | −1.771 | 0.625 |
| FGR | V2 | 0.720 (0.688–0.753) | 0.073 | 0.043 | −1.750 | 0.898 |
| FGR | V3 | 0.811 (0.782–0.839) | 0.129 | 0.039 | −1.829 | 0.948 |

AUROC, area under the receiver operating characteristic curve; AUPRC, area under the precision–recall curve.

**Supplementary Table S8. Sensitivity of trajectory-group comparisons to conformal confidence level**

| Outcome | Confidence | Comparison (group 1 − group 0) | Group 1 n/events (%) | Group 0 n/events (%) | Crude RD, pp (95% CI) | Adjusted RD, pp (95% CI) |
|---|---|---|---|---|---|---|
| GDM | 80% | Persistent high − emerging high | 1,172/156 (13.3%) | 548/43 (7.8%) | +5.46 (+2.62 to +8.47) | +0.78 (−3.37 to +4.77) |
| GDM | 80% | Stable low − resolved-to-low | 3,005/40 (1.3%) | 1,143/33 (2.9%) | −1.56 (−2.67 to −0.56) | −0.94 (−2.11 to +0.10) |
| GDM | 90% | Persistent high − emerging high | 552/96 (17.4%) | 341/36 (10.6%) | +6.83 (+2.43 to +11.47) | +1.09 (−4.36 to +6.15) |
| GDM | 90% | Stable low − resolved-to-low | 1,440/11 (0.8%) | 1,302/28 (2.2%) | −1.39 (−2.35 to −0.55) | −1.11 (−2.06 to +0.00) |
| PE | 80% | Persistent high − emerging high | 1,053/166 (15.8%) | 820/80 (9.8%) | +6.01 (+3.14 to +8.93) | +2.49 (−1.08 to +6.06) |
| PE | 80% | Stable low − resolved-to-low | 2,309/49 (2.1%) | 1,262/40 (3.2%) | −1.05 (−2.15 to +0.07) | −0.78 (−2.05 to +0.38) |
| PE | 90% | Persistent high − emerging high | 462/84 (18.2%) | 503/70 (13.9%) | +4.27 (−0.09 to +8.93) | −0.47 (−5.35 to +4.47) |
| PE | 90% | Stable low − resolved-to-low | 1,046/19 (1.8%) | 1,197/29 (2.4%) | −0.61 (−1.74 to +0.49) | −0.65 (−1.78 to +0.46) |
| PTB | 80% | Persistent high − emerging high | 873/151 (17.3%) | 898/117 (13.0%) | +4.27 (+1.13 to +7.53) | −0.62 (−3.86 to +2.96) |
| PTB | 80% | Stable low − resolved-to-low | 1,486/50 (3.4%) | 1,646/68 (4.1%) | −0.77 (−2.10 to +0.53) | −0.07 (−1.49 to +1.27) |
| PTB | 90% | Persistent high − emerging high | 386/89 (23.1%) | 547/95 (17.4%) | +5.69 (+0.26 to +10.87) | +0.53 (−4.94 to +5.82) |
| PTB | 90% | Stable low − resolved-to-low | 655/14 (2.1%) | 1,320/48 (3.6%) | −1.50 (−2.90 to +0.05) | −1.05 (−2.60 to +0.67) |
| LGA | 80% | Persistent high − emerging high | 777/230 (29.6%) | 1,134/295 (26.0%) | +3.59 (−0.42 to +7.63) | −3.97 (−8.00 to +0.15) |
| LGA | 80% | Stable low − resolved-to-low | 1,739/54 (3.1%) | 2,068/91 (4.4%) | −1.30 (−2.47 to −0.05) | −0.53 (−1.74 to +0.80) |
| LGA | 90% | Persistent high − emerging high | 310/114 (36.8%) | 722/243 (33.7%) | +3.12 (−2.96 to +9.12) | −6.07 (−12.17 to +0.37) |
| LGA | 90% | Stable low − resolved-to-low | 743/18 (2.4%) | 2,184/71 (3.3%) | −0.83 (−2.12 to +0.55) | −0.03 (−1.54 to +1.54) |
| FGR | 80% | Persistent high − emerging high | 509/39 (7.7%) | 1,155/91 (7.9%) | −0.22 (−2.95 to +2.49) | −2.76 (−5.26 to −0.14) |
| FGR | 80% | Stable low − resolved-to-low | 1,557/7 (0.4%) | 2,854/23 (0.8%) | −0.36 (−0.82 to +0.15) | −0.15 (−0.67 to +0.46) |
| FGR | 90% | Persistent high − emerging high | 186/17 (9.1%) | 721/79 (11.0%) | −1.82 (−6.58 to +3.20) | −3.80 (−8.09 to +0.92) |
| FGR | 90% | Stable low − resolved-to-low | 603/3 (0.5%) | 3,301/19 (0.6%) | −0.08 (−0.61 to +0.65) | +0.27 (−0.43 to +1.37) |

Risk differences are group 1 minus group 0 and are shown in percentage points (pp). The 90% rows correspond to the primary analysis and the 80% rows to the sensitivity analysis. Adjusted estimates used logistic regression of outcome on trajectory group and a four-degree-of-freedom natural cubic spline of Visit 3 predicted probability, with g-computation standardized over the combined sample of the two groups in each contrast. Confidence intervals used 2,000 participant-level bootstrap resamples.

**Supplementary Table S9. Trajectory-group composition across conformal confidence levels**

**Panel A. GDM**

| Confidence | Trajectory group | n | % | Events | Event rate | Complete-trajectory denominator |
|---|---|---|---|---|---|---|
| 80% | Persistent high | 1,172 | 13.6% | 156 | 13.3% | 8,623 |
| 80% | Emerging high | 548 | 6.4% | 43 | 7.8% | 8,623 |
| 80% | Intermittent high | 138 | 1.6% | 5 | 3.6% | 8,623 |
| 80% | Stable low | 3,005 | 34.8% | 40 | 1.3% | 8,623 |
| 80% | Resolved-to-low | 1,143 | 13.3% | 33 | 2.9% | 8,623 |
| 80% | Prior high to low | 70 | 0.8% | 0 | 0.0% | 8,623 |
| 80% | Review-ending | 2,547 | 29.5% | 95 | 3.7% | 8,623 |
| 90% | Persistent high | 552 | 6.4% | 96 | 17.4% | 8,623 |
| 90% | Emerging high | 341 | 4.0% | 36 | 10.6% | 8,623 |
| 90% | Intermittent high | 60 | 0.7% | 8 | 13.3% | 8,623 |
| 90% | Stable low | 1,440 | 16.7% | 11 | 0.8% | 8,623 |
| 90% | Resolved-to-low | 1,302 | 15.1% | 28 | 2.2% | 8,623 |
| 90% | Prior high to low | 2 | 0.0% | 0 | 0.0% | 8,623 |
| 90% | Review-ending | 4,926 | 57.1% | 193 | 3.9% | 8,623 |

**Panel B. PE**

| Confidence | Trajectory group | n | % | Events | Event rate | Complete-trajectory denominator |
|---|---|---|---|---|---|---|
| 80% | Persistent high | 1,053 | 11.4% | 166 | 15.8% | 9,262 |
| 80% | Emerging high | 820 | 8.9% | 80 | 9.8% | 9,262 |
| 80% | Intermittent high | 133 | 1.4% | 13 | 9.8% | 9,262 |
| 80% | Stable low | 2,309 | 24.9% | 49 | 2.1% | 9,262 |
| 80% | Resolved-to-low | 1,262 | 13.6% | 40 | 3.2% | 9,262 |
| 80% | Prior high to low | 121 | 1.3% | 4 | 3.3% | 9,262 |
| 80% | Review-ending | 3,564 | 38.5% | 173 | 4.9% | 9,262 |
| 90% | Persistent high | 462 | 5.0% | 84 | 18.2% | 9,262 |
| 90% | Emerging high | 503 | 5.4% | 70 | 13.9% | 9,262 |
| 90% | Intermittent high | 64 | 0.7% | 11 | 17.2% | 9,262 |
| 90% | Stable low | 1,046 | 11.3% | 19 | 1.8% | 9,262 |
| 90% | Resolved-to-low | 1,197 | 12.9% | 29 | 2.4% | 9,262 |
| 90% | Prior high to low | 7 | 0.1% | 0 | 0.0% | 9,262 |
| 90% | Review-ending | 5,983 | 64.6% | 312 | 5.2% | 9,262 |

**Panel C. PTB**

| Confidence | Trajectory group | n | % | Events | Event rate | Complete-trajectory denominator |
|---|---|---|---|---|---|---|
| 80% | Persistent high | 873 | 10.0% | 151 | 17.3% | 8,748 |
| 80% | Emerging high | 898 | 10.3% | 117 | 13.0% | 8,748 |
| 80% | Intermittent high | 140 | 1.6% | 19 | 13.6% | 8,748 |
| 80% | Stable low | 1,486 | 17.0% | 50 | 3.4% | 8,748 |
| 80% | Resolved-to-low | 1,646 | 18.8% | 68 | 4.1% | 8,748 |
| 80% | Prior high to low | 195 | 2.2% | 9 | 4.6% | 8,748 |
| 80% | Review-ending | 3,510 | 40.1% | 253 | 7.2% | 8,748 |
| 90% | Persistent high | 386 | 4.4% | 89 | 23.1% | 8,748 |
| 90% | Emerging high | 547 | 6.3% | 95 | 17.4% | 8,748 |
| 90% | Intermittent high | 68 | 0.8% | 8 | 11.8% | 8,748 |
| 90% | Stable low | 655 | 7.5% | 14 | 2.1% | 8,748 |
| 90% | Resolved-to-low | 1,320 | 15.1% | 48 | 3.6% | 8,748 |
| 90% | Prior high to low | 12 | 0.1% | 0 | 0.0% | 8,748 |
| 90% | Review-ending | 5,760 | 65.8% | 413 | 7.2% | 8,748 |

**Panel D. LGA**

| Confidence | Trajectory group | n | % | Events | Event rate | Complete-trajectory denominator |
|---|---|---|---|---|---|---|
| 80% | Persistent high | 777 | 9.2% | 230 | 29.6% | 8,453 |
| 80% | Emerging high | 1,134 | 13.4% | 295 | 26.0% | 8,453 |
| 80% | Intermittent high | 117 | 1.4% | 25 | 21.4% | 8,453 |
| 80% | Stable low | 1,739 | 20.6% | 54 | 3.1% | 8,453 |
| 80% | Resolved-to-low | 2,068 | 24.5% | 91 | 4.4% | 8,453 |
| 80% | Prior high to low | 515 | 6.1% | 30 | 5.8% | 8,453 |
| 80% | Review-ending | 2,103 | 24.9% | 237 | 11.3% | 8,453 |
| 90% | Persistent high | 310 | 3.7% | 114 | 36.8% | 8,453 |
| 90% | Emerging high | 722 | 8.5% | 243 | 33.7% | 8,453 |
| 90% | Intermittent high | 52 | 0.6% | 16 | 30.8% | 8,453 |
| 90% | Stable low | 743 | 8.8% | 18 | 2.4% | 8,453 |
| 90% | Resolved-to-low | 2,184 | 25.8% | 71 | 3.3% | 8,453 |
| 90% | Prior high to low | 83 | 1.0% | 2 | 2.4% | 8,453 |
| 90% | Review-ending | 4,359 | 51.6% | 498 | 11.4% | 8,453 |

**Panel E. FGR**

| Confidence | Trajectory group | n | % | Events | Event rate | Complete-trajectory denominator |
|---|---|---|---|---|---|---|
| 80% | Persistent high | 509 | 5.9% | 39 | 7.7% | 8,666 |
| 80% | Emerging high | 1,155 | 13.3% | 91 | 7.9% | 8,666 |
| 80% | Intermittent high | 159 | 1.8% | 9 | 5.7% | 8,666 |
| 80% | Stable low | 1,557 | 18.0% | 7 | 0.4% | 8,666 |
| 80% | Resolved-to-low | 2,854 | 32.9% | 23 | 0.8% | 8,666 |
| 80% | Prior high to low | 997 | 11.5% | 14 | 1.4% | 8,666 |
| 80% | Review-ending | 1,435 | 16.6% | 27 | 1.9% | 8,666 |
| 90% | Persistent high | 186 | 2.1% | 17 | 9.1% | 8,666 |
| 90% | Emerging high | 721 | 8.3% | 79 | 11.0% | 8,666 |
| 90% | Intermittent high | 67 | 0.8% | 5 | 7.5% | 8,666 |
| 90% | Stable low | 603 | 7.0% | 3 | 0.5% | 8,666 |
| 90% | Resolved-to-low | 3,301 | 38.1% | 19 | 0.6% | 8,666 |
| 90% | Prior high to low | 257 | 3.0% | 3 | 1.2% | 8,666 |
| 90% | Review-ending | 3,531 | 40.7% | 84 | 2.4% | 8,666 |

Percentages use the complete three-visit trajectory denominator for the corresponding outcome and confidence level. The seven trajectory groups follow the definitions in Supplementary Table S3.

**Supplementary Table S10. Visit-specific empirical coverage overall and within observed outcome classes**

**Panel A. GDM**

| Confidence | Visit | Subset | n | Covered | Empirical coverage (95% CI) | Empty sets | Without a prediction at this visit, n |
|---|---|---|---|---|---|---|---|
| 80% | Visit 1 | Overall | 8,720 | 6,996 | 80.2% (79.4 to 81.1) | 0 (0.0%) | 0 |
| 80% | Visit 1 | Class 0 | 8,344 | 6,695 | 80.2% (79.4 to 81.1) | 0 (0.0%) | 0 |
| 80% | Visit 1 | Class 1 | 376 | 301 | 80.1% (75.7 to 83.8) | 0 (0.0%) | 0 |
| 80% | Visit 2 | Overall | 8,720 | 7,028 | 80.6% (79.8 to 81.4) | 0 (0.0%) | 0 |
| 80% | Visit 2 | Class 0 | 8,344 | 6,730 | 80.7% (79.8 to 81.5) | 0 (0.0%) | 0 |
| 80% | Visit 2 | Class 1 | 376 | 298 | 79.3% (74.9 to 83.0) | 0 (0.0%) | 0 |
| 80% | Visit 3 | Overall | 8,623 | 6,896 | 80.0% (79.1 to 80.8) | 0 (0.0%) | 97 |
| 80% | Visit 3 | Class 0 | 8,251 | 6,597 | 80.0% (79.1 to 80.8) | 0 (0.0%) | 97 |
| 80% | Visit 3 | Class 1 | 372 | 299 | 80.4% (76.0 to 84.1) | 0 (0.0%) | 97 |
| 90% | Visit 1 | Overall | 8,720 | 7,894 | 90.5% (89.9 to 91.1) | 0 (0.0%) | 0 |
| 90% | Visit 1 | Class 0 | 8,344 | 7,553 | 90.5% (89.9 to 91.1) | 0 (0.0%) | 0 |
| 90% | Visit 1 | Class 1 | 376 | 341 | 90.7% (87.3 to 93.2) | 0 (0.0%) | 0 |
| 90% | Visit 2 | Overall | 8,720 | 7,847 | 90.0% (89.3 to 90.6) | 0 (0.0%) | 0 |
| 90% | Visit 2 | Class 0 | 8,344 | 7,498 | 89.9% (89.2 to 90.5) | 0 (0.0%) | 0 |
| 90% | Visit 2 | Class 1 | 376 | 349 | 92.8% (89.8 to 95.0) | 0 (0.0%) | 0 |
| 90% | Visit 3 | Overall | 8,623 | 7,771 | 90.1% (89.5 to 90.7) | 0 (0.0%) | 97 |
| 90% | Visit 3 | Class 0 | 8,251 | 7,438 | 90.1% (89.5 to 90.8) | 0 (0.0%) | 97 |
| 90% | Visit 3 | Class 1 | 372 | 333 | 89.5% (86.0 to 92.2) | 0 (0.0%) | 97 |

**Panel B. PE**

| Confidence | Visit | Subset | n | Covered | Empirical coverage (95% CI) | Empty sets | Without a prediction at this visit, n |
|---|---|---|---|---|---|---|---|
| 80% | Visit 1 | Overall | 9,289 | 7,473 | 80.4% (79.6 to 81.2) | 0 (0.0%) | 0 |
| 80% | Visit 1 | Class 0 | 8,737 | 7,028 | 80.4% (79.6 to 81.3) | 0 (0.0%) | 0 |
| 80% | Visit 1 | Class 1 | 552 | 445 | 80.6% (77.1 to 83.7) | 0 (0.0%) | 0 |
| 80% | Visit 2 | Overall | 9,289 | 7,400 | 79.7% (78.8 to 80.5) | 0 (0.0%) | 0 |
| 80% | Visit 2 | Class 0 | 8,737 | 6,986 | 80.0% (79.1 to 80.8) | 0 (0.0%) | 0 |
| 80% | Visit 2 | Class 1 | 552 | 414 | 75.0% (71.2 to 78.4) | 0 (0.0%) | 0 |
| 80% | Visit 3 | Overall | 9,262 | 7,422 | 80.1% (79.3 to 80.9) | 0 (0.0%) | 27 |
| 80% | Visit 3 | Class 0 | 8,737 | 6,990 | 80.0% (79.2 to 80.8) | 0 (0.0%) | 27 |
| 80% | Visit 3 | Class 1 | 525 | 432 | 82.3% (78.8 to 85.3) | 0 (0.0%) | 27 |
| 90% | Visit 1 | Overall | 9,289 | 8,375 | 90.2% (89.5 to 90.7) | 0 (0.0%) | 0 |
| 90% | Visit 1 | Class 0 | 8,737 | 7,879 | 90.2% (89.5 to 90.8) | 0 (0.0%) | 0 |
| 90% | Visit 1 | Class 1 | 552 | 496 | 89.9% (87.1 to 92.1) | 0 (0.0%) | 0 |
| 90% | Visit 2 | Overall | 9,289 | 8,344 | 89.8% (89.2 to 90.4) | 0 (0.0%) | 0 |
| 90% | Visit 2 | Class 0 | 8,737 | 7,860 | 90.0% (89.3 to 90.6) | 0 (0.0%) | 0 |
| 90% | Visit 2 | Class 1 | 552 | 484 | 87.7% (84.7 to 90.2) | 0 (0.0%) | 0 |
| 90% | Visit 3 | Overall | 9,262 | 8,350 | 90.2% (89.5 to 90.7) | 0 (0.0%) | 27 |
| 90% | Visit 3 | Class 0 | 8,737 | 7,873 | 90.1% (89.5 to 90.7) | 0 (0.0%) | 27 |
| 90% | Visit 3 | Class 1 | 525 | 477 | 90.9% (88.1 to 93.0) | 0 (0.0%) | 27 |

**Panel C. PTB**

| Confidence | Visit | Subset | n | Covered | Empirical coverage (95% CI) | Empty sets | Without a prediction at this visit, n |
|---|---|---|---|---|---|---|---|
| 80% | Visit 1 | Overall | 8,853 | 7,044 | 79.6% (78.7 to 80.4) | 0 (0.0%) | 0 |
| 80% | Visit 1 | Class 0 | 8,081 | 6,432 | 79.6% (78.7 to 80.5) | 0 (0.0%) | 0 |
| 80% | Visit 1 | Class 1 | 772 | 612 | 79.3% (76.3 to 82.0) | 0 (0.0%) | 0 |
| 80% | Visit 2 | Overall | 8,853 | 7,062 | 79.8% (78.9 to 80.6) | 0 (0.0%) | 0 |
| 80% | Visit 2 | Class 0 | 8,081 | 6,452 | 79.8% (79.0 to 80.7) | 0 (0.0%) | 0 |
| 80% | Visit 2 | Class 1 | 772 | 610 | 79.0% (76.0 to 81.7) | 0 (0.0%) | 0 |
| 80% | Visit 3 | Overall | 8,748 | 6,997 | 80.0% (79.1 to 80.8) | 0 (0.0%) | 105 |
| 80% | Visit 3 | Class 0 | 8,081 | 6,457 | 79.9% (79.0 to 80.8) | 0 (0.0%) | 105 |
| 80% | Visit 3 | Class 1 | 667 | 540 | 81.0% (77.8 to 83.8) | 0 (0.0%) | 105 |
| 90% | Visit 1 | Overall | 8,853 | 7,950 | 89.8% (89.2 to 90.4) | 0 (0.0%) | 0 |
| 90% | Visit 1 | Class 0 | 8,081 | 7,252 | 89.7% (89.1 to 90.4) | 0 (0.0%) | 0 |
| 90% | Visit 1 | Class 1 | 772 | 698 | 90.4% (88.1 to 92.3) | 0 (0.0%) | 0 |
| 90% | Visit 2 | Overall | 8,853 | 7,964 | 90.0% (89.3 to 90.6) | 0 (0.0%) | 0 |
| 90% | Visit 2 | Class 0 | 8,081 | 7,260 | 89.8% (89.2 to 90.5) | 0 (0.0%) | 0 |
| 90% | Visit 2 | Class 1 | 772 | 704 | 91.2% (89.0 to 93.0) | 0 (0.0%) | 0 |
| 90% | Visit 3 | Overall | 8,748 | 7,877 | 90.0% (89.4 to 90.7) | 0 (0.0%) | 105 |
| 90% | Visit 3 | Class 0 | 8,081 | 7,272 | 90.0% (89.3 to 90.6) | 0 (0.0%) | 105 |
| 90% | Visit 3 | Class 1 | 667 | 605 | 90.7% (88.3 to 92.7) | 0 (0.0%) | 105 |

**Panel D. LGA**

| Confidence | Visit | Subset | n | Covered | Empirical coverage (95% CI) | Empty sets | Without a prediction at this visit, n |
|---|---|---|---|---|---|---|---|
| 80% | Visit 1 | Overall | 8,509 | 6,817 | 80.1% (79.3 to 80.9) | 0 (0.0%) | 0 |
| 80% | Visit 1 | Class 0 | 7,541 | 6,037 | 80.1% (79.1 to 80.9) | 0 (0.0%) | 0 |
| 80% | Visit 1 | Class 1 | 968 | 780 | 80.6% (78.0 to 82.9) | 0 (0.0%) | 0 |
| 80% | Visit 2 | Overall | 8,509 | 6,809 | 80.0% (79.2 to 80.9) | 0 (0.0%) | 0 |
| 80% | Visit 2 | Class 0 | 7,541 | 6,037 | 80.1% (79.1 to 80.9) | 0 (0.0%) | 0 |
| 80% | Visit 2 | Class 1 | 968 | 772 | 79.8% (77.1 to 82.2) | 0 (0.0%) | 0 |
| 80% | Visit 3 | Overall | 8,453 | 6,800 | 80.4% (79.6 to 81.3) | 0 (0.0%) | 56 |
| 80% | Visit 3 | Class 0 | 7,491 | 6,013 | 80.3% (79.4 to 81.2) | 0 (0.0%) | 56 |
| 80% | Visit 3 | Class 1 | 962 | 787 | 81.8% (79.2 to 84.1) | 0 (0.0%) | 56 |
| 90% | Visit 1 | Overall | 8,509 | 7,673 | 90.2% (89.5 to 90.8) | 0 (0.0%) | 0 |
| 90% | Visit 1 | Class 0 | 7,541 | 6,786 | 90.0% (89.3 to 90.6) | 0 (0.0%) | 0 |
| 90% | Visit 1 | Class 1 | 968 | 887 | 91.6% (89.7 to 93.2) | 0 (0.0%) | 0 |
| 90% | Visit 2 | Overall | 8,509 | 7,706 | 90.6% (89.9 to 91.2) | 0 (0.0%) | 0 |
| 90% | Visit 2 | Class 0 | 7,541 | 6,816 | 90.4% (89.7 to 91.0) | 0 (0.0%) | 0 |
| 90% | Visit 2 | Class 1 | 968 | 890 | 91.9% (90.1 to 93.5) | 0 (0.0%) | 0 |
| 90% | Visit 3 | Overall | 8,453 | 7,651 | 90.5% (89.9 to 91.1) | 0 (0.0%) | 56 |
| 90% | Visit 3 | Class 0 | 7,491 | 6,780 | 90.5% (89.8 to 91.2) | 0 (0.0%) | 56 |
| 90% | Visit 3 | Class 1 | 962 | 871 | 90.5% (88.5 to 92.2) | 0 (0.0%) | 56 |

**Panel E. FGR**

| Confidence | Visit | Subset | n | Covered | Empirical coverage (95% CI) | Empty sets | Without a prediction at this visit, n |
|---|---|---|---|---|---|---|---|
| 80% | Visit 1 | Overall | 8,787 | 7,010 | 79.8% (78.9 to 80.6) | 0 (0.0%) | 0 |
| 80% | Visit 1 | Class 0 | 8,542 | 6,810 | 79.7% (78.9 to 80.6) | 0 (0.0%) | 0 |
| 80% | Visit 1 | Class 1 | 245 | 200 | 81.6% (76.3 to 86.0) | 0 (0.0%) | 0 |
| 80% | Visit 2 | Overall | 8,787 | 6,971 | 79.3% (78.5 to 80.2) | 0 (0.0%) | 0 |
| 80% | Visit 2 | Class 0 | 8,542 | 6,767 | 79.2% (78.3 to 80.1) | 0 (0.0%) | 0 |
| 80% | Visit 2 | Class 1 | 245 | 204 | 83.3% (78.1 to 87.4) | 0 (0.0%) | 0 |
| 80% | Visit 3 | Overall | 8,666 | 6,938 | 80.1% (79.2 to 80.9) | 0 (0.0%) | 121 |
| 80% | Visit 3 | Class 0 | 8,456 | 6,772 | 80.1% (79.2 to 80.9) | 0 (0.0%) | 121 |
| 80% | Visit 3 | Class 1 | 210 | 166 | 79.0% (73.0 to 84.0) | 0 (0.0%) | 121 |
| 90% | Visit 1 | Overall | 8,787 | 7,902 | 89.9% (89.3 to 90.5) | 0 (0.0%) | 0 |
| 90% | Visit 1 | Class 0 | 8,542 | 7,677 | 89.9% (89.2 to 90.5) | 0 (0.0%) | 0 |
| 90% | Visit 1 | Class 1 | 245 | 225 | 91.8% (87.7 to 94.7) | 0 (0.0%) | 0 |
| 90% | Visit 2 | Overall | 8,787 | 7,901 | 89.9% (89.3 to 90.5) | 0 (0.0%) | 0 |
| 90% | Visit 2 | Class 0 | 8,542 | 7,674 | 89.8% (89.2 to 90.5) | 0 (0.0%) | 0 |
| 90% | Visit 2 | Class 1 | 245 | 227 | 92.7% (88.7 to 95.3) | 0 (0.0%) | 0 |
| 90% | Visit 3 | Overall | 8,666 | 7,768 | 89.6% (89.0 to 90.3) | 0 (0.0%) | 121 |
| 90% | Visit 3 | Class 0 | 8,456 | 7,583 | 89.7% (89.0 to 90.3) | 0 (0.0%) | 121 |
| 90% | Visit 3 | Class 1 | 210 | 185 | 88.1% (83.0 to 91.8) | 0 (0.0%) | 121 |

Coverage was counted when the prediction set contained the observed class. Empty sets were counted as not covered; no empty sets occurred in these outputs. Visits for which no evaluable conformal state was produced were excluded from the visit-specific denominator. Confidence intervals are Wilson intervals.

**Supplementary Table S11. TRIPOD+AI checklist**

Reporting was assessed against TRIPOD+AI for the current manuscript and Supplementary Information.

| TRIPOD+AI item | Reporting in the present study | Location / current status |
|---|---|---|
| 1. Title | The title identifies longitudinal prediction of multiple adverse pregnancy outcomes and the antenatal context. | Title. |
| 2. Abstract | The abstract reports the cohort, five outcomes, three antenatal visits, conformal prediction approach, principal longitudinal findings, and empirical coverage. | Abstract. |
| 3a. Healthcare context and rationale | The Introduction describes repeated antenatal risk assessment, limitations of isolated time-point predictions, and the rationale for examining prediction histories and longitudinal uncertainty. | Introduction. |
| 3b. Target population, intended purpose and users | The target population is nulliparous women with singleton pregnancies. Models were investigated for longitudinal antenatal risk stratification rather than as an implemented clinical decision-support system. | Introduction and Methods. |
| 3c. Health inequalities | No dedicated health-inequality analysis was performed. | Not performed. |
| 4. Objectives | Objectives include development of outcome-specific models at three visits, evaluation of whether prediction history adds information beyond the Visit 3 predicted probability, cross-outcome prediction changes, and longitudinal conformal coverage. | End of Introduction. |
| 5a. Data source and representativeness | Existing data from the prospective multicenter nuMoM2b cohort were used. | Methods, Study population. |
| 5b. Data collection dates | Participants were enrolled between September 2010 and September 2013. | Methods, Study population. |
| 6a. Setting | nuMoM2b recruited participants at eight US clinical centers. | Methods, Study population. |
| 6b. Eligibility criteria | Nulliparous women with singleton pregnancies were included; outcome-specific eligibility and Visit 3 landmark restrictions are described. | Methods; Supplementary Table S2. |

| TRIPOD+AI item | Reporting in the present study | Location / current status |
|---|---|---|
| 6c. Treatments received | No treatment was assigned as part of the present observational prediction study. | Not applicable as a study-assigned intervention; treatments received in routine care were not summarized for this analysis. |
| 7. Data preprocessing and quality checking | Predictors were restricted by visit availability and ≤20% missingness. Continuous variables were imputed by five-nearest-neighbor imputation and scaled using training-set 5th and 95th percentiles; categorical missingness was encoded as a separate category before one-hot encoding. All preprocessing was fitted within the relevant training data and applied unchanged to evaluation data. | Methods. |
| 8a. Outcomes and prediction horizon | GDM, PE, PTB, LGA and FGR definitions and ascertainment are reported; GDM and PE followed nuMoM2b study definitions. Predictions were generated at three gestational windows. | Methods, Adverse pregnancy outcomes; Supplementary Method S1. |
| 8b. Subjective outcome assessment | FGR was based on the nuMoM2b maternal chart-abstraction item recording suspected antenatal fetal growth restriction before delivery (CMAE08). Additional clinical details were abstracted when FGR was suspected. | Methods, Adverse pregnancy outcomes. Abstractor qualifications not reported in the source documentation. |
| 8c. Blinding of outcome assessment | Blinding of outcome ascertainment to predictor information was documented for GDM, PE, PTB and LGA. Corresponding information was not available for the FGR chart-abstraction variable. | Source documentation for GDM, PE, PTB and LGA; FGR blinding status not documented. |
| 9a. Initial predictor selection | Candidate predictors were derived from previous nuMoM2b studies, restricted by availability at each visit and ≤20% missingness; the same visit-specific candidate set was used across outcomes. | Methods, Prediction models and conformal prediction. |
| 9b. Predictor definitions and timing | Predictor domains and visit availability are provided. | Supplementary Table S1. |
| 9c. Subjectively assessed predictors | Some source variables involved operator-dependent measurements, including ultrasonography and Doppler. Gestational-age ultrasonography was performed by certified sonographers. | Methods, Adverse pregnancy outcomes. Assessor details for other measurements not reported in the source documentation. |

| TRIPOD+AI item | Reporting in the present study | Location / current status |
|---|---|---|
| 10. Sample size | This secondary analysis used data from all 9,289 participants who consented to data sharing; outcome-specific analysis sets are reported. | Methods, Study population and data source; Supplementary Table S2. |
| 11. Missing data | Predictors with >20% missingness were excluded. Remaining continuous missing values were imputed using five-nearest-neighbor imputation (k=5), and missing categorical values were treated as a separate category within the relevant training data. Missingness for the reported baseline characteristics is provided in Table 1. | Methods; Table 1. |
| 12a. Data use and partitioning | Subject-level stratified 10-fold cross-validation was used. For conformal prediction, 20% of each outer training portion was reserved for calibration and the model was fitted on the remaining proper-training subset. Held-out participants were not used for model fitting or calibration. | Methods, Prediction models and conformal prediction. |
| 12b. Predictor handling | Continuous predictors were truncated and scaled using the training-set 5th and 95th percentiles. Missing categorical values were encoded as a separate category before one-hot encoding. Fitted transformations were applied unchanged to evaluation data. | Methods. |
| 12c. Model type, model building, tuning and internal validation | Separate elastic-net logistic regression models were fitted for each outcome and visit. Hyperparameters were selected by fivefold internal cross-validation using AUROC, over l1_ratio {0.1, 0.3, 0.5, 0.7, 0.9} and C {0.001, 0.01, 0.1, 0.5, 1}; performance was evaluated using subject-level 10-fold cross-validation. | Methods. |
| 12d. Heterogeneity across clusters | Models were not developed using center-specific effects and performance heterogeneity across the eight clinical centers was not evaluated. | Not evaluated. |
| 12e. Model-performance measures | AUROC was the primary discrimination measure. AUPRC, Brier score and calibration measures were reported as secondary performance measures. Conformal coverage was assessed separately. | Methods; Supplementary Table S7. |

| TRIPOD+AI item | Reporting in the present study | Location / current status |
|---|---|---|
| 12f–g. Model updating / evaluation of an existing model | No external evaluation or updating of a previously developed fixed prediction model was performed. | Not applicable. |
| 13. Class imbalance | During model fitting, the positive-class weight was set to the square root of the negative-to-positive sample ratio in the relevant training data; the negative-class weight was 1. | Methods. |
| 14. Fairness | No dedicated fairness-aware modeling or subgroup fairness analysis was performed. | Not performed. |
| 15. Model output and classification | Models generated event probabilities. Mondrian split conformal prediction converted these into prediction sets represented as low, review, high or empty states using class-specific calibration thresholds. The primary confidence level was 90%. | Methods; Supplementary Method S2. |
| 16. Development versus evaluation data | Model development and internal evaluation were conducted within the same cohort using held-out cross-validation; no separate external evaluation dataset was used. | Methods. |
| 17. Ethical approval | The present analysis used data from participants who consented to data sharing; NICHD and Eindhoven University of Technology ethical approvals are reported. | Methods, Study population and data source. |
| 18a. Funding | No specific funding was received for the present study. | No specific funding. |
| 18b. Conflicts of interest | The authors declare no competing interests. | Competing interests statement. |
| 18c. Protocol | No separate study-specific protocol was developed for the present secondary analysis. The parent nuMoM2b cohort design and data-collection procedures have been described previously. | Methods, Study population and data source; published nuMoM2b methods paper (reference 19). |
| 18d. Registration | The present secondary analysis was not prospectively registered. | Not registered. |
| 18e. Data sharing | nuMoM2b data are available through the NICHD Data and Specimen Hub (DASH), subject to approval by the DASH Data Access Committee. The original and processed analytical datasets are not redistributed with the code. | Data availability statement. |

| TRIPOD+AI item | Reporting in the present study | Location / current status |
|---|---|---|
| 18f. Code sharing | Code for model development, prediction, conformal calibration and downstream analyses is available at https://github.com/yanqi-tue/conformal_prediction_yq; the controlled-access nuMoM2b data must be obtained independently through NICHD DASH. | Code availability statement; software versions are reported in Methods. |
| 19. Patient and public involvement | No patient or public involvement was undertaken in the design, conduct, reporting, or dissemination planning of the present secondary analysis. | No patient or public involvement in the present secondary analysis. |
| 20a. Participant flow | Overall and outcome-specific analysis-set sizes are reported, including Visit 3 landmark restrictions. | Results; Supplementary Table S2. |
| 20b. Participant characteristics | Baseline characteristics of the full study cohort are reported, including maternal age, BMI, race/ethnicity, education, selected medical history and Visit 1 blood pressure. | Table 1. |
| 20c. Development versus evaluation characteristics | Internal cross-validation rather than a separate evaluation cohort was used. | Not directly applicable as an external-evaluation comparison. |
| 21. Participants/events in each analysis | Outcome-specific denominators and trajectory group sizes are reported. | Supplementary Tables S2–S3. |
| 22. Full model specification | The model class, training procedure and analysis code are reported, but fitted coefficients or model objects sufficient to reproduce the existing predictions directly are not provided. | Methods and Code availability statement; fitted-model objects are not provided. |
| 23a. Model performance with uncertainty | AUROC estimates with 95% confidence intervals are reported across visits; AUPRC, Brier score and calibration measures are reported in Supplementary Table S7. | Results; Supplementary Table S7. Subgroup performance/fairness evaluation was not performed. |
| 23b. Cluster heterogeneity | Performance heterogeneity across clinical centers was not examined. | Not performed. |
| 24. Model updating | No external model updating was performed. | Not applicable. |
| 25. Interpretation | Results are interpreted in relation to prediction history, the Visit 3 predicted probability, uncertainty, cross-outcome dependence and longitudinal coverage, with comparison to previous work. | Discussion. |

| TRIPOD+AI item | Reporting in the present study | Location / current status |
|---|---|---|
| 26. Limitations | Limitations include use of a single antenatal cohort without external validation, three visit windows, FGR ascertainment as a recorded antenatal suspicion, overlap of GDM and PE diagnoses with Visit 3, restriction of Visit 3 analyses to pregnancies continuing to 30+0 weeks, and absence of evaluation of clinical consequences. | Discussion. |
| 27a. Handling unavailable/poor-quality inputs in implementation | The study did not evaluate prospective implementation or handling of unavailable inputs at deployment. | Not evaluated in the present study. |
| 27b. User interaction/expertise | No clinical interface or deployment workflow was evaluated. | Not applicable to the current research implementation. |
| 27c. Future applicability/generalizability | The Discussion identifies limitations of the present cohort and the need for longitudinal calibration methods and further evaluation. | Discussion. |

**Supplementary Figures**

**Supplementary Figure S1. Sensitivity of the persistent-high versus emerging-high comparison to model-support restriction and matching**

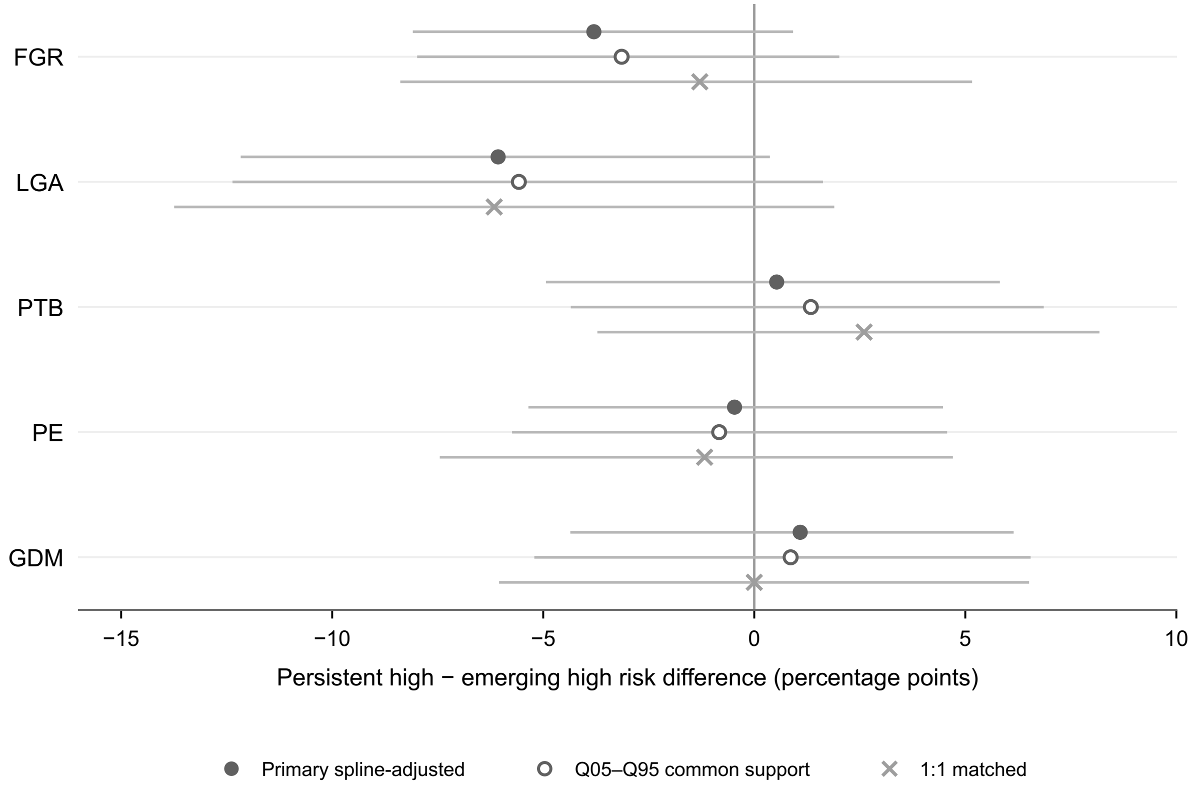


Risk differences are persistent high minus emerging high. Filled circles show the primary spline-adjusted estimates using the full comparison sample; open circles show the same spline adjustment after restriction to the common-support interval (overlap of the 5th–95th percentile ranges) of Visit 3 predicted probability; crosses show 1:1 nearest-neighbor matched estimates on the logit of Visit 3 predicted probability. Horizontal lines indicate 95% confidence intervals. The common-support analyses used 2,000 bootstrap resamples and the matching analysis used 500 bootstrap resamples.
